# An emerging TIMER-2C framework for addressing barriers to research culture and productivity among local healthcare providers in the Middle East and sub-Saharan Africa: a qualitative study and modified Delphi approach

**DOI:** 10.1101/2025.06.20.25327944

**Authors:** Beryne Odeny, Dorothy Mangale, Dhananjaya Sharma, Mariam Balogun, Elizabeth Bukusi, Fatima Suleman, Miliard Derbew, Juliet Iwelunmor, Ermyas Birru, Isaac Che Ngang, Akila Anandarajah, Vaishnavi Mamillapalle, Wael Almahmeed

## Abstract

**Background:** Contributions from healthcare professionals in the Middle East and North Africa (MENA) and sub-Saharan Africa (SSA) to global health research remain disproportionately low. we conceptualized the TIMER-2C framework as emerging approach to capture the multilevel barriers and facilitators shaping research productivity among local healthcare professionals in MENA and SSA.

**Methods:** We conducted qualitative interviews with a modified Delphi approach comprising two rounds of interviewing to: 1) identify barriers and facilitators, and 2) establish consensus on the most prominent factors influencing research productivity. We purposively sampled 17 established and early career health researchers from Jordan, Qatar, Egypt, Lebanon, North Sudan, South Sudan, Nigeria, Ghana, Kenya, Uganda, Ethiopia, South Africa and India. Interview guides were grounded in the “Capacity Strengthening of Health Research in Conflict” and the “Health Equity Implementation” frameworks. We explored barriers, facilitators, and recommendations for increasing research output. Transcripts were managed in ATLAS.ti and thematically analyzed to identify emerging themes.

**Results:** Barriers and facilitators to research productivity occurred at individual, institutional, national, and global levels. Key barriers included limited time, funding, mentorship, and research training; institutional emphasis on clinical and teaching duties over research; lack of national investment and supportive policies; and global inequities in funding, publishing, and collaboration. Facilitators included personal motivation, protected time, access to training and mentorship, institutional support for research, national funding strategies, and equitable global partnerships that value research contributions from local healthcare professionals. We identified six interlinked core themes and one overarching theme from which we coined the **TIMER-2C** conceptual framework to represent: 1) **<u>T</u>**ime, 2) **<u>I</u>**nstability, Interest & Infrastructure, 3) **<u>M</u>**oney & Resources, 4) **<u>E</u>**xpertise & Experience, 5) **<u>R</u>**ecognition, and 6) constructive **<u>C</u>**ollaboration, which are elements that promote a positive research **<u>C</u>**ulture (2C stands for Collaboration and Culture) that builds research capacity and creates a sustainable pipeline of researchers.

**Conclusion:** We identified research culture as an overarching influence on research contributions from MENA and SSA. We unveil an emerging conceptual framework, the *TIMER-2C framework*, that depicts and explains the relationship between factors associated with research productivity. This framework with its seven constructs can be a useful tool for guiding and/or evaluating local and global strategies to increase research output. We recommend further use of this framework to guide the design, implementation and evaluation of multi-pronged strategies to enhance research contributions from healthcare professionals in MENA, SSA, and other similar contexts.

## INTRODUCTION

### Underrepresentation of local healthcare professional researchers in health research

Health research is a cornerstone of productive health systems and communities. However, there remains anunderrepresentation of research led by local healthcare professionals/clinicians who are native to the region and/or working within local institutions—in the Middle East and North Africa (MENA) and sub-Saharan Africa (SSA). This gap calls for further exploration into how healthcare professionals and institutions in these regions value and support research, including fostering enabling policies, institutional norms, and societal attitudes that uphold research. While some progress has been made in recent decades, research output from MENA and SSA continues to lag behind that of other regions, despite bearing a disproportionate share of the global disease burden.^1,2^

### Regional disparities in funding and priority setting and gender inequities in research

Ideally, health research output should align with national or local health priorities, but many low- and middle-income countries (LMICs) in MENA and SSA heavily depend on external research and funding, often from high-income countries (HICs) which may be contextually misaligned with priorities in LMICs.^3^ This mismatch presents two main challenges: 1) a disconnect between the disease burden and research funding and thus low research output, and 2) the superimposing of external research priorities and evidence that may not align with local realities, resulting in suboptimal health care outcomes.

Ignacio Atal and colleagues., in a 2018 study, found a mismatch between research effort, i.e., volume of randomized controlled trials, and the distribution of major causes of burden, especially infectious diseases and neonatal disorders in LMICs.^4^ In the MENA region, a review found gaps in Global Health Capacity Building initiatives to improve health workforce performance with limited coverage of regional health priorities such as conflict, emergencies, and non-communicable diseases.^5^ Gender disparities further compound these challenges. In the MENA and SSA, women constitute 72% of the health workforce; their representation declines to only 35% among medical doctors and 25% in top healthcare leadership positions.^6,7^ Women are also underrepresented in research (30-40% of researchers) and are less likely than men to publish over the course of their academic careers.^8–10^ Further, men publish between 11 and 51% more than women, with a widening gap over time.^11^ In SSA, the ratio of female to male health researchers was reported as 9:17 in health research institutions, highlighting persistent gender imbalance in the research productivity among healthcare professionals.^12^

### Research challenges and vulnerability

Researchers within SSA and the MENA have low access to research funding and infrastructure which hinders research capacity greatly in these regions. Despite the significant financial resources in some MENA countries, a 2020 study reported under-investment in biomedical research, shortage of well-trained researchers, and limited government support for research.^13^ Moreover, while collaboration with HIC institutions is common, researchers from SSA and MENA institution are often underrepresented in leadership prominence and authorship of joint publications.^14^ These challenges leave MENA and SSA vulnerable to parachute or extractive science, where research is conducted by external researchers with limited local involvement or benefit.^15^

We sought to understand determinants of local research productivity in the MENA and SSA, and broadly in low income countries (LICs), and middle income countries (MICs). By examining contextual factors and existing solutions implemented by individuals and institutions, strategic actions can be identified to empower local researchers to increase their contributions to the global health research landscape. The specific objectives of this inquiry were to identify the main barriers and facilitators influencing the conduct of high-quality health research by local healthcare professionals in the MENA and SSA regions.

### Conceptual Framework

This study was guided by a modified conceptual framework for strengthening the capacity of health research in conflict regions in the MENA by El Achi et.al.^16^ El Achi et al.’s framework highlights the structural layers influencing research capacity—from national priorities, sociopolitical forces, funding, and institutional infrastructure to individual skills, mentorship, and gender roles that shape research productivity in conflict-affected MENA settings. This framework was developed to address the gap in available research capacity strengthening frameworks that fail to target fragile, conflict-affected contexts. *We added the global level to the multiple layers*.

We also incorporated the Health Equity Implementation Framework to address structural inequities often overlooked in capacity building models.^17^ We emphasized equity determinants at each level, including societal influences (political and economic), organizational and individual contexts, day-to-day encounters and interactions, historical marginalization, and power imbalance. We depict the role of facilitation strategies such as relationship building in supporting implementation of programs to promote research. Finally, monitoring and evaluation is necessary to ensure appropriate and consistent outcomes. Mapping these frameworks onto each other enables examination of how barriers and facilitators to equitable research capacity building are shaped by power dynamics and inequities across levels. For example, institutional infrastructure (El Achi) maps onto organizational context, individual interactions and facilitation (Woodward), while individual-level factors such as time and skills (El Achi) map onto individual interactions and facilitation (Woodward). We capture both the multilevel determinants and equity dimensions that affect who participates and whose priorities are advanced in research capacity building.

## MATERIALS AND METHODS

We conducted qualitative interviews and a modified Delphi approach. Qualitative interview guides with prompts were created based on the domains and constructs of the integrated conceptual framework. The study was conducted between January and March 2024 and was in line with the Consolidated criteria for Reporting Qualitative Research (COREQ) guidelines, a checklist for the reporting of qualitative studies^18^ (appendix 1).

### Participant selection

We used purposive sampling, through a snowballing approach, to identify and email key informants from HICs and LMICs. We recruited leading or emerging researchers involved in authorship, mentorship, and publishing from 14 countries in MENA, SSA (English-speaking countries), and Asia (HICs: Qatar, United Arab Emirates, MICs: Egypt, Jordan, South Africa and India, LICs: Lebanon, North Sudan, South Sudan, Nigeria, Ghana, Kenya, Uganda, Ethiopia,). Of the 36 individuals contacted via email, 17 verbally consented to participate—8 from MENA, 8 from SSA and 1 from Asia. The low participation rate was due to competing professional obligations and limited availability which hindered scheduling of interviews. Some were caught up in volatile environments and had to seek refuge in other countries, thus were unable to keep their scheduled appointments.

The emails of emerging researchers were obtained through a snowballing approach, where initial participants identified through professional networks referred eligible researchers. Table 1 providers the characteristics of the participants. **Inclusion criteria** included local healthcare professionals, history of involvement in health research in MENA, SSA, or other LMIC settings; experience in publishing; and willingness to participate in virtual interviews. **Exclusion criteria** included external researchers and those who have conducted research exclusively in HICs.

**Table 1:**
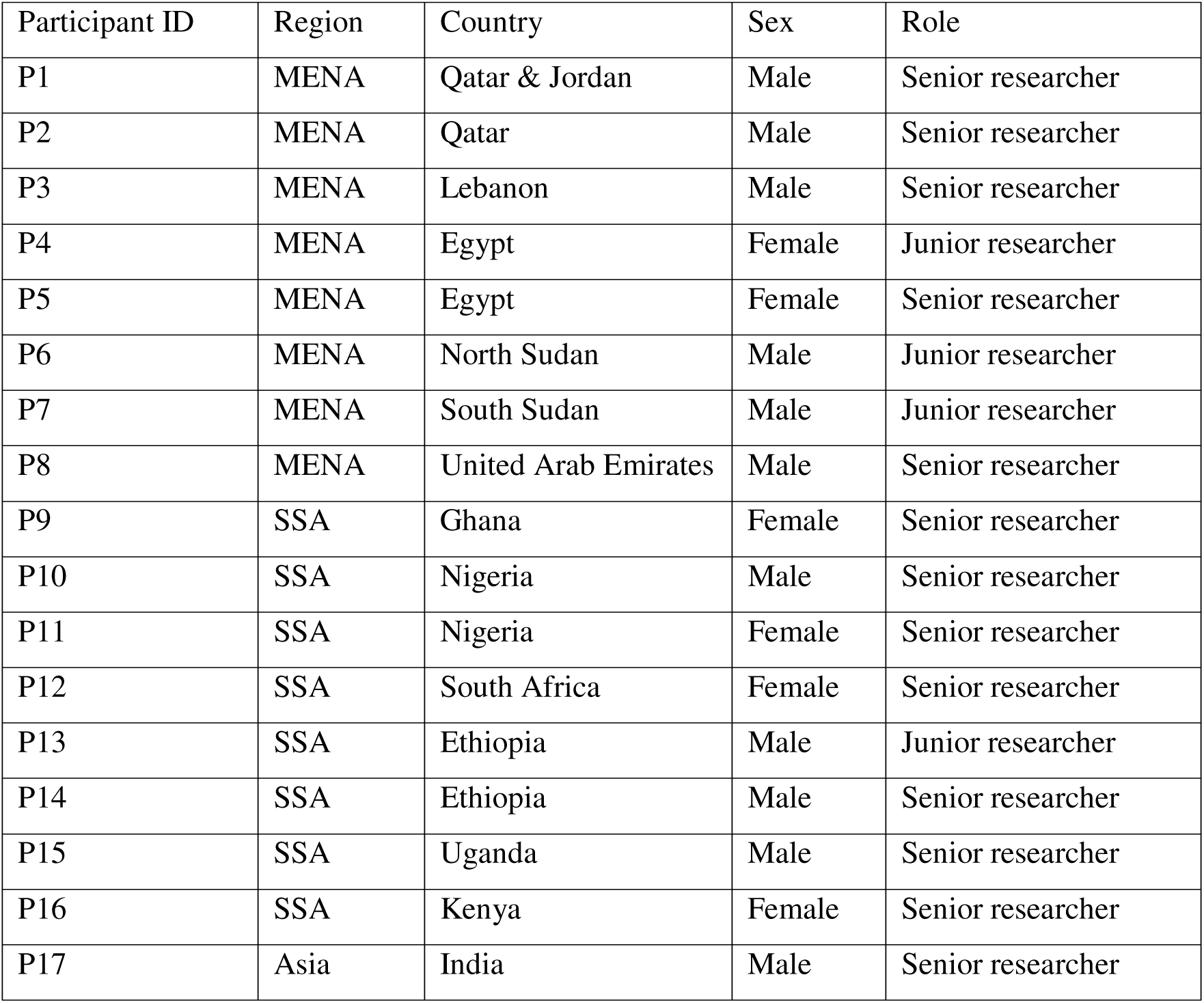
Participant characteristics.

| Participant ID | Region | Country | Sex | Role |
| --- | --- | --- | --- | --- |
| P1 | MENA | Qatar & Jordan | Male | Senior researcher |
| P2 | MENA | Qatar | Male | Senior researcher |
| P3 | MENA | Lebanon | Male | Senior researcher |
| P4 | MENA | Egypt | Female | Junior researcher |
| P5 | MENA | Egypt | Female | Senior researcher |
| P6 | MENA | North Sudan | Male | Junior researcher |
| P7 | MENA | South Sudan | Male | Junior researcher |
| P8 | MENA | United Arab Emirates | Male | Senior researcher |
| P9 | SSA | Ghana | Female | Senior researcher |
| P10 | SSA | Nigeria | Male | Senior researcher |
| P11 | SSA | Nigeria | Female | Senior researcher |
| P12 | SSA | South Africa | Female | Senior researcher |
| P13 | SSA | Ethiopia | Male | Junior researcher |
| P14 | SSA | Ethiopia | Male | Senior researcher |
| P15 | SSA | Uganda | Male | Senior researcher |
| P16 | SSA | Kenya | Female | Senior researcher |
| P17 | Asia | India | Male | Senior researcher |

**Table 2:**
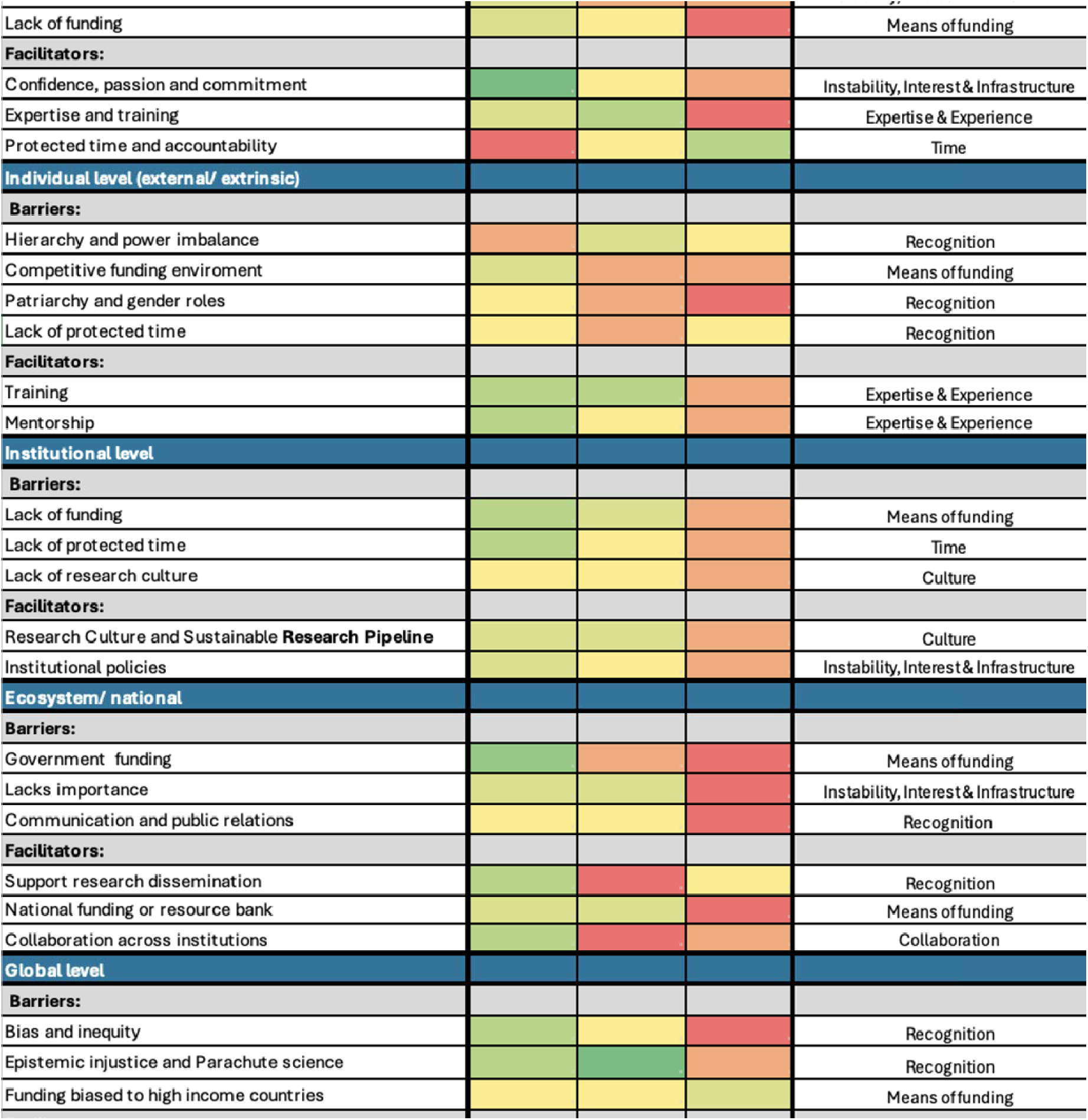
Modified Delphi Approach Ranking Heat Map at Multiple Levels Table 1 footnote: A factor was considered a top priority if more than 5 participants ranked it as their number one. Factors ranked among the top three by 4–5 participants were given preeminence and prioritized. Factors ranked as top three by 3 or fewer participants were assigned lower importance.

|  |  |  |  |  |
| --- | --- | --- | --- | --- |
| Lack of funding |  |  |  | Means of funding |
| **Facilitators:** |  |  |  |  |
| Confidence, passion and commitment |  |  |  | Instability, Interest & Infrastructure |
| Expertise and training |  |  |  | Expertise & Experience |
| Protected time and accountability |  |  |  | Time |
| **Individual level (external/ extrinsic)** |  |  |  |  |
| **Barriers:** |  |  |  |  |
| Hierarchy and power imbalance |  |  |  | Recognition |
| Competitive funding environment |  |  |  | Means of funding |
| Patriarchy and gender roles |  |  |  | Recognition |
| Lack of protected time |  |  |  | Recognition |
| **Facilitators:** |  |  |  |  |
| Training |  |  |  | Expertise & Experience |
| Mentorship |  |  |  | Expertise & Experience |
| **Institutional level** |  |  |  |  |
| **Barriers:** |  |  |  |  |
| Lack of funding |  |  |  | Means of funding |
| Lack of protected time |  |  |  | Time |
| Lack of research culture |  |  |  | Culture |
| **Facilitators:** |  |  |  |  |
| Research Culture and Sustainable Research Pipeline |  |  |  | Culture |
| Institutional policies |  |  |  | Instability, Interest & Infrastructure |
| **Ecosystem/ national** |  |  |  |  |
| **Barriers:** |  |  |  |  |
| Government funding |  |  |  | Means of funding |
| Lacks importance |  |  |  | Instability, Interest & Infrastructure |
| Communication and public relations |  |  |  | Recognition |
| **Facilitators:** |  |  |  |  |
| Support research dissemination |  |  |  | Recognition |
| National funding or resource bank |  |  |  | Means of funding |
| Collaboration across institutions |  |  |  | Collaboration |
| **Global level** |  |  |  |  |
| **Barriers:** |  |  |  |  |
| Bias and inequity |  |  |  | Recognition |
| Epistemic injustice and Parachute science |  |  |  | Recognition |
| Funding biased to high income countries |  |  |  | Means of funding |

### Data collection

The interview guide was calibrated during the initial interview to ensure that questions were clear and elicited appropriate responses. Interviews were scheduled at the key informants’ availability. Prior to the interviews, the research team introduced themselves and shared the objectives for this study. The study team (BO and DM) conducted two rounds of interviews over Zoom. The first round of interviews focused on gathering experiences and perspectives on barriers, facilitators, and recommendations for increasing research output by local healthcare professionals in the MENA and SSA. In the second round of interviews, a modified Delphi process was used to identify and rank top factors. This was a modified Delphi in that participants provided rankings in the presence of the study team and only two rounds of interviewing were conducted.

Interviews took approximately 45 minutes and were audio-recorded with consent for accurate transcription of data. Only the interviewers and key informants were present during the interviews. Notes were taken during and after the interview. Interviewers debriefed on key topics that emerged from the in-depth interviews. By the 17th interview, we determined that data saturation was achieved, because no new topics or themes emerged; consequently, we ceased efforts to contact additional interviewees. All interviews were conducted in English, and no translation was required. To ensure transcript quality, each transcript was reviewed by the interviewer after transcription to verify accuracy and completeness.

### Data analysis

Transcripts were uploaded to ATLAS.ti for coding and thematic analysis. We employed a hybrid deductive-inductive thematic analysis approach. We analyzed the qualitative data using deductive coding (based on a priori codes from the conceptual framework) and inductive coding to capture emerging themes in the data. Initial codes were informed by the conceptual frameworks that were then refined to produce a unified coding scheme with code groups and individual codes. Codes were grouped into broader themes, stratified by level (individual, institutional, national, global) and mapped to key domains (barriers, facilitators, recommendations).

A modified Delphi approach was applied with 16 of the participants in two rounds. A consensus was established for key factors across five levels: individual, interpersonal (external individual), institutional, national/ecosystem, and global. A factor was considered a top priority if more than 5 participants ranked it as their number one. Factors ranked among the top three by 4–5 participants were given preeminence and prioritized. Factors ranked as top three by 3 or fewer participants were assigned lower importance. A heat map was created to summarize the most frequently identified barriers and facilitators.

### Research Team and Reflexivity

The research team consisted of the principal investigator (BO) – a medical doctor and assistant professor of surgery at Washington University in St. Louis, and co-investigator (DM) – a postdoctoral fellow in the department of medicine at Washington University in St. Louis, who coded and established intercoder agreement. Both BO and DM are women of Kenyan descent, trained and worked in the Kenya both doing programmatic and research work that heavily involved global collaborations prior to their overseas training in public health, and qualitative and quantitative methods. Both hold PhDs in Global health and Implementation Science. The team’s positionality as researchers originally from SSA but currently based at a high-income country (HIC) institution, adds both insider and outsider knowledge into the dynamics of research productivity. The senior author, WA, is a practicing clinician and researcher in the United Arab Emirates, where he is originally from. This was acknowledged during data collection and interpretation.

### Human subjects research statement

Ethics approval was not sought prior to this work, as it was considered minimal risk; however, retrospective IRB review determined that project would have qualified for exempt status. This study solely involved academic discussions with globally distributed researchers to identify barriers and facilitators of research productivity in the MENA and SSA regions. Informed consent was obtained verbally from each participant prior to the commencement of Zoom discussions.

## RESULTS

The study included 17 participants from MENA, SSA, and Asia. The sample included both senior (n=13) and junior (n=4) researchers, with a majority being senior level. There were 10 males and 7 females with different levels of research experience.

The thematic analysis yielded six overarching determinants influencing research productivity: 1) Time; 2) Instability, Interest and Infrastructure; 3) Means of funding; 4) Experience/Expertise; and 5) Recognition. Two additional cross-cutting themes—constructive Collaboration and Culture—emerged consistently across interviews and had influence on research at multiple levels. These seven themes were organized into a new framework called TIMER-2C, an acronym derived from the initials of the five main factors (T, I, M, E, R) and the two cross-cutting themes (2C for constructive Collaboration and Culture).

## THEMES

### 1. Time

Time emerged as a multidimensional factor manifesting primarily as: a) lack of protected time for research by healthcare professionals, b) the delayed nature or long duration between research and impact (with the need for patience and long-term commitment to reap the rewards of research), and c) the need for sustainability or continuity of research to build a research identity and culture. The ability to maneuver these dimensions of time greatly dictates a healthcare professional’s or institution’s willingness to participate in meaningful and transformative research in LMICs.

“It’s a mentorship thing. You are not going to grow a good researcher in a day. Research is a commitment for a period of time – mentorship-attitude, mentorship-growth, encouraging people to truly mentor and build the next generation.” [SSA senior female researcher]

**At the individual level,** some healthcare professionals would opt for the immediate benefits of clinical work over the benefits of research which take a long time to realize. They also cite struggling to balance competing demands of health care provision, teaching and research. For some, focus on one means a loss of the other and thus no middle ground. For professionals who contemplate research, there is a general feeling of exhaustion and therefore an inability to lend oneself to research—on top of clinical care—as this would compromise patient care and sacrifice clinical income.

“They cannot graduate without producing a scholarly project… The problem, however, after they graduate is the residency programs. Where primarily service, as you know, takes over everything else.” [MENA senior female researcher]

“We are overwhelmed… so you are really just barely surviving, just trying to stay from drowning in the service delivery work… You are actually overwhelmed… I would have burnt out completely if I had not left… it wasn’t humanly sustainable.” [SSA senior male researcher]

**At the interpersonal and institutional level**, participants identified a systemic failure to support protected time for research time through funding, institutional policy, or workload adjustments. Institutional priorities often skewed toward clinical service delivery, teaching, and administrative tasks leaving little room for research.

“You will be very surprised many universities did not even consider my application and they were very frank. Some of them say, ‘… really your career path is research, what we need are teachers, and you don’t have any teaching experience.’ It was indicated to me very explicitly. So, why? because there are huge teaching loads, universities are not sufficiently funded to allow time for research for the faculty. So, they teach a lot of courses. and that means there is no room for research over there or very, very limited research outside.” [MENA senior male researcher]

“Again, the reasons are more structural as compared to just educational in facilitating time commitment, service versus academia. All of those things are—I think they’re not helping in creating more physician scientists.” [MENA senior female researcher]

**At the national level,** There are exceptional and high-functioning institutions that build and support research capacity in MENA and SSA like the Aga Khan University, American University of Beirut, and Makerere University, among others. In MENA, research is not always published for political and national security reasons. On the other hand, several medical schools or universities were perceived to lack conducive research environments. The perceived lack of a research culture within some institutions was seen to contribute to weak policies that don’t protect research time, underdeveloped research support systems, and poor administrative infrastructure for training programs, mentorship support, and career opportunities for researchers. For instance, we learned that research professors often have burdensome teaching or clinical loads and often have challenges being absorbed into institutions or being compensated as research faculty.

“It becomes a policy level at the governmental level. Or if it’s a big institution, at the institutional level… somebody ultimately has to take care of patients. Do you have enough physicians to take care of patients and are you—you get stuck there. Unless there is enough funds to be able to recruit enough clinicians to take care of patients so that they’ll be able to pay the researchers, then that could change.” [MENA senior female researcher]

### 2. Instability, Interest and Infrastructure

Political and economic instability are major barriers to research production. In unstable environments, some participants pointed out that basic safety and survival take precedence over research interests. Without infrastructure, accessible data and transparent systems, researchers face significant barriers in advancing impactful research. Participants demonstrated how research interest is linked to investment in research infrastructure like research administrative offices, regulatory bodies, supportive institutional policies, and training resources.

**At the individual level,** healthcare professionals often deprioritize research in favor of providing emergency care and clinical services needed for survival in times of *instability*. This directly impacts individual interest in pursuing research. Nevertheless, *interest* in research was not uncommon in fragile and unstable contexts. Two researchers from MENA reported that their interest was sparked during their formative years in high school and fueled their passion, sacrifice and commitment to research. Passion was identified as a driving force that enabled researchers to make the most of limited opportunities. However, without support and motivation—especially in the absence of distinct research career paths or outcomes—many healthcare professionals disengaged from research.

“Based on my experience setting up a clinical study across five countries, I think one of the other unique issues in the Middle East is also the environment or the political—let me put it very straight, the political instability… when I started my study, Syria was one of the collaborating centers and they had to drop because the war in Syria had started at that time.” [MENA senior female researcher]

“If I have opportunities but there is no passion, if there is no commitment then we are wasting time.” [SSA senior female researcher]

“She drove me through, and she gave me the passion, she supported me in every step.” [MENA junior female researcher]

“At a certain point, you have to make a concerted effort to improve it. My writing has improved, because I read, I observed, I changed but some people are stuck …” [SSA senior female researcher]

**Interpersonal** interaction and exposure to research through mentorship and peer networks – when supported with reliable *infrastructure* – can promote *interest* in research but are often severely disrupted by political and institutional *instability*. Without a stable research environment, even strong peer or mentor relationships may not be enough to sustain interest in research.

“I talked to a female scientist… Before the war, she traveled and left her baby son … she had to leave everything and go to the UAE to work remotely from there to help her students. Even when she was in Sudan, she had challenges to get the material needed for her research. Because of the economic sanctions on Sudan, she couldn’t get it.” [MENA senior female researcher]

“There could be very good mentors that could be very good sponsors but if you yourself are not driven, and don’t take advantage of that mentorship and sponsorship… you may become a researcher but that’s not the ideal situation or you might not get to the potential that you might want to get to.” [SSA senior female researcher]

**At the institutional level**, there was mention of a lack of *interest* and a general feeling that “institutions don’t care.” We learned of researchers who were frustrated by the system, especially those who had international research experience. They noted that having protected time for research is not enough if the institution does not have an interest in advancing research. Participants emphasized the need for *infrastructure* such as administrative support, regulatory structures for ethics approval, supportive institutional policies, internet broadband, and training resources. In fact, research professors often have challenges being absorbed into institutions or being compensated as research faculty. In the midst of *instability*, institutions often cannot provide the necessary environment for research due to disrupted operations.

“There are huge teaching loads, universities are not sufficiently funded to allow time for research for the faculty. [MENA senior male researcher]

“These are things that our institutions miss actually and the value of research. Many supervisors don’t actually know the value of research or appreciate the value of research. So, how are they going to teach this or give it to the candidates?” [MENA junior female researcher]

“It’s an individual issue because it’s also created by the system. Because the system, in most academia here, values how many publications versus really the value of the publications.” [MENA senior female researcher]

**At the national level**, *interest* and commitment is a huge driver of research. In some settings interest is perceived to be low and public institutions often face limited *infrastructure* and funding, making them doubly disadvantaged when compared to their HIC counterparts. *Instability* also discourages national research planning. In certain countries or regions that had robust research, there was suppression of research findings or dissemination for sociopolitical reasons. Findings related to mortality and stigmatizing issues were suppressed and censored. In some countries, there was minimal emphasis on research and low appreciation for its utility, thus low curiosity and passion for it at institutional and national levels.

“We have a lot of good policies that we do as a country, like there is a national research ethnics group… We have a database where if you want a part do research in a province better get a proposal this province give you feedback, they give you all of that. So that system is nice.” [SSA senior female researcher]

“Do research for preparedness for 20 years. What you mean preparedness? I’m already in the middle of a problem… What we are dealing with is maybe catchup game because of funding and because of that stability in many parts. All my devotion … is basically to take care of our people. Prevent deaths, prevent hunger—so it’s very hard to do research on how to reduce obesity. Nobody will give you money to how to increase physical activity …” [MENA senior male researcher]

“…the situation doesn’t allow you to go and research while you are really struggling to pay your physicians to take care of patients or to keep your hospitals running.” [MENA senior male researcher]

**At the global level,** sociopolitical and economic *instability* within countries in the MENA and SSA further exacerbates these challenges by deterring the establishment of robust global research ecosystems.

“The issue is funding, stability, being able to have access. I cannot get any data in Pakistan. Nobody can get data from Pakistan. It’s guarded like a secret. You can’t get mortality data from the Gulf because they’re afraid to share their—in many countries, there are a limitation for what you can do. Sexual transmitted diseases, you don’t talk about… Alcohol, you don’t talk about. … we have the data, and they collect it, but they don’t share it and they don’t talk about…” [MENA senior male researcher]

“They do a lot of surveys. They do a lot of surveillance system. You can debate the quality as much as you want. Many of them don’t release the data; that’s the problem. You don’t hear about it because data is power in the region. Many people don’t like to share the data because they may lose some of that power. I wouldn’t say there is no research. There is a lot of research. There is a lot of good researchers.” [MENA senior male researcher]

### 3. Means of funding research

Access to equitable funding was a major facilitator of research across all levels; yet, participants consistently highlighted how local and global disparities in funding allocation and management hinder research productivity in LMICs.

**At the individual level,** participants cited limited access to resources to support research, training costs or fees to obtain things like the internet for virtual learning and publishing. In one setting, funding for student researchers’ fieldwork had been embezzled or redirected to other initiatives. Funding issues were associated with loss of talent through brain-drain and many skilled clinical researchers seeking opportunities outside their countries.

Participants noted that the lack of funding restricts how much time they can spend generating ideas or writing scientific articles, thereby limiting their contribution to various fields. Several participants explained that they had to maintain several professional appointments, mostly clinical, including running their private practices, to generate funds for their research. Self-funded researchers often face the challenge of balancing fundraising efforts with their actual research activities, which can lead to insufficient time for conducting research.

“The main issue in my country mainly is the funding of research… that prevents us from doing high quality research, we self-fund about 90% of our projects… so we pay a lot of expenses on research for example the transportation, the consumables, all the data forms… but the passion drives us more than the money of course. [MENA junior female researcher]

“I can tell you, I train people… They can do research. They’re hired by WHO, and they leave the next day after we train them. If there is funding and there is money, and you could do what you want to do. If I can do what I’m doing here in my country, I wouldn’t be here for a minute; I’d go back…” [MENA senior male researcher]

“I paid for my own research with my own personal funds that I generated from my clinical practice. It is the interest in research that drives you…” [Asia senior male researcher]

That’s what helped me and the passion. If you have to passion towards something, then you can do it.” [MENA junior male researcher]

One opposing view to the money factor was that funding specifically is not necessary to conduct research. Lack of funding was perceived as an opportunity to fend for ones research, eliminate donor-dependency, and find resourceful ways for institutions to match funding. One participant mentioned their practice was to do “pure science/ research” free from contamination of donor-funded agendas – instead, they harnessed local resources to answer locally pertinent questions.

“So you have to find local solutions. None of my publications has received any funding: Zero funding, and we used the resource which is commonly available to everyone—our knowledge.” [ASIA senior male researcher]

“The thing is that they also come with their agendas. Agenda is not necessarily a bad thing, but they also come with their agendas… Again, there are others have had foundation grants where honestly, they have no agenda except we have to fund you research” [SSA senior male researcher]

**At the interpersonal level,** there was mention of resource sharing, teamwork, avoiding waste, and supporting fellow researchers with leftover materials to help alleviate funding gaps.

“We share the resources, you know you work as a team in every project. We cherish working as a team, for example, if someone has some remaining materials, they can share with their colleague to complete the work and so on.” [MENA junior female researcher]

**At the institutional level,** researchers in LMICs often lack the supportive infrastructure available to their counterparts in HICs, hindering their ability to effectively apply for large-scale funding opportunities. Poor stewardship of financial resources has led to disappointment due to a lack of structures to absorb or manage grant funds, and in some instances, administrative structures are biased or corrupt. There were instances in which institutional investment and funding was high, and researchers felt supported.

“There was funding allocated for students to do their field work…and do the data collection, every student got some money to do that… We have been hearing different stories that there is corruption, money goes somewhere else unfortunately.” [MENA junior male researcher]

“… has quite enlightened leadership… They decided to invest in science and in education, in fact I work here, and it is called education city. Literally it is a city for education science and research… Fortunately we have good access to funding… I am basically enjoying my career… We have had extremely high productivity; I can tell you for sure if I was in the USA I could not have had this kind of success.” [MENA senior male researcher]

**At the national level,** funding is needed for training, infrastructure including administration, human resources, regulatory processes and protected research time. Health research budgets in LMICs are almost non-existent and strong economies like Qatar and South Africa were found to have substantial health financing budgets that catered for healthcare research. Uganda too had some local funds set apart for health research. Compared to participants from SSA, participants from MENA were more likely to use their personal funds or source local funds for research. In other settings, corruption and embezzlement of research funds was a barrier to research engagement.

“ Money, time hits all of it. So, money to me explains why you can have 10 hours, so many publications or grants…. they have the culture and the funding… But even that aside, just the fact that the funding is there… at the government level, we don’t fund ” [SSA senior female researcher]

“Challenges are, the first one, funding. They don’t have access, like I do, to NIH as a funder. That’s one. The funding from these agencies is very limited. Sometimes, it’s conditional-only partnership Sometimes, it’s not. If it’s not at NIH, it’s really peanuts. It’s not much. That’s one. The second, the local funding. The funding from the government for research is not that great. Many of their universities—except certain universities in the region—a professor is fully paid and doesn’t have to—he or she—hustle for funds.” [MENA senior male researcher]

“We have a funding authority in Egypt, its name is the STDF, The science and technology development fund… it contributes to public health.” [MENA junior female researcher]

**At the global level,** lack of funding emerged as predominant determinant of the slow growth of research in MENA and the SSA region. Participants highlighted that most funding for research comes from HICs. There are very few examples of LMICs in SSA and MENA internally funding their research. LMICs have limited budgets for health which cascades to minimal funding for health research. Challenges accessing limited external funding in a competitive global environment, stringent requirements by granting institutions, and donor agendas that were misaligned with local research agendas were cited. There were recommendations to locally co-fund research by matching funded grants to encourage researchers.

“It’s like the aid question again, if we had our own funding for health systems and our health programs, we would have our own targets. We would be less disadvantaged in determining the direction of our health programs …but when you have the external funding, it is not bad – but the extent to which it funds research to a large proportion, it changes agendas, it changes the way of thinking, it changes the value system. I can be in an international collaboration, and more respect is given to the White people . I will be sitting there wondering, who am I even in this thing? Nobody even respects what I am saying…” [SSA senior female researcher]

“Encourage them, it could even be co-funding by the way… if someone for example would apply and they get some kind of funding we match it, something that is pretty common in the united states… So in this way also we support their successes.” [MENA senior male researcher]

### 4. Expertise and Experience

Beyond access to protected time, interest (passion and commitment), infrastructure, stability, and funding, training, mentorship and sponsorship, and research opportunities are needed to propel individuals in their research career paths and promote a research culture.

**At the individual level**, training was the link that connected an individual’s passion with their preconceived research idea. Many successful researchers sought training beyond what they had in their formal clinical coursework and benefited from supportive mentorship. In some settings – particularly in SSA – healthcare professionals sometimes were not well-trained. Some participants expressed that the lack of confidence experienced by researchers contributed to a demotivated mindset, hindering their pursuit of training and knowledge in research. Additionally, the language barrier – particularly in non-English speaking regions – posed a major challenge, that impeded the development of technical research skills, especially scientific writing.

“…since third year medical school I got into research and what helped me was Youtube, online courses etc. For me that was the only way to learn. I faced a lot of struggles… I was staying at the university until after midnight just to have internet access to download the course” [MENA junior male researcher]

“What we have found out with our students who have done structured research training – they were more likely to publish in residency, more likely to get research awards in residency, and more likely to continue in academia.” [MENA senior female researcher]

There is a language barrier to some extent… people have the perception that it all has to be in English and they don’t feel comfortable…” [MENA senior female researcher]

**At the interpersonal level,** co-occurring with the lack of expertise was the limited access to, or in some cases, poor quality—or variable availability—of mentorship for emerging researchers. Many participants had trouble finding mentors, while others were subject to mentors who did not always have the required expertise. In other instances, the power dynamics embedded in mentor-mentee relationships fostered mistreatment/ abuse of students and early career researchers, leading to transactional exchanges lacking authentic relationships and supportive environments.

“Our system and culture are so authoritarian that we don’t do very good mentoring. It’s only a few people who mentor, a lot of professors who mentor people are doing it begrudgingly because it’s part of the expectation and they terrorize a lot of people in that system. So, mentorship is not great, and people do not want to go into those fields.” [SSA senior female researcher]

“The institution needs extensive training and better communication between mentors, supervisors and candidates. Most of the candidates actually work on their own without monitoring from their supervisors,…” [MENA junior female researcher].

” …mentorship, mentorship-attitude, mentorship-growth. encouraging people to truly mentor and build the next generation requires a little bit of selflessness… you have to be willing to allow someone else to overtake you… How do we grow that culture, in a world where everybody wants to be top dog.” [SSA senior female researcher]

“It’s a group that was created by some recent graduate from Egypt they are helping people to learn about systematic reviews…, all the process, selecting the idea…, screening and writing the manuscript.” [MENA junior male researcher]

**At the institutional level,** some participants indicated that healthcare professionals are trained primarily for service and not for research. Others noted the absence of training in research writing in universities, saying they were, “not trained to write,” while others pointed out that in the early stages of their involvement in research, nobody explained aspects like journal rankings or how to select journals for publication. This was particularly the case in SSA.

“We are not training people in how to communicate and disseminate research especially in the written form. Oral is all very good and its excellent… but a lot of what goes to really change the research world needs to be written, needs to be published, needs to be in this peer reviewed format.” [SSA senior male researcher]

“I would support building more STEM schools in the region because I believe that it starts with younger people. People in the region need to be told earlier in life about science and technology… I think maybe we need to go earlier in the stage rather than supporting researchers after graduation.” [MENA senior female researcher]

“We don’t train people to be researchers to start with. We don’t encourage curiosity…, the child who is curious that’s the child who is “naughty.” So, we don’t encourage curiosity, we don’t encourage experimentation, so we kill research very early on because to be a researcher, and to think research, you need to think out of the box.” [SSA senior female researcher]

**At the national and global levels,** there is a lot of research expertise in various parts of MENA and SSA as illustrated by world renown health researchers and studies from these regions. Unfortunately, many trained healthcare professionals leave these countries for better opportunities. Emerging researchers struggle with lack research training, mentorship, and sponsorship. This was cited as one of the reasons why parachute research took root in LMICs as the expertise and experiences came from external researchers, whereas indigenous/ local researchers were undermined. Another cited barrier with regard to expertise and experience was “brain drain” as trained health professionals exit their countries in search for better paying opportunities or further training and do not to return for fear of frustration and lack of job satisfaction. One participant indicated that if the resources and opportunities were available in their home country, they would return to establish their research labs.

“There’s a lot of research. Whether it is published or not, that’s totally different… If you take Iran, for example, there is a lot of research universities, publications, studies. In Pakistan, there is a lot of research. Most of the information we have about enteric diseases comes from one of the labs that is run by the Aga Khan University… the American University of Beirut. [MENA senior male researcher]

“Ideas of the projects were coming from high income countries… in low-income countries we just become collectors, and we would not be mentioned in articles…” [SSA senior male researcher]

“Brain drain. All the people who do good research—look at Pakistan, Aga Khan—I can give you examples at AUB as well. Aga Khan University, the person who did an amazing job … are recruited by the Gates Foundation, by the Sick Children in Canada…” [MENA senior male researcher]

### 5. Recognition

Recognition is a multifaceted determinant that captures research and researcher acknowledgment both locally and globally. This domain encapsulates self-recognition, institutional recognition, societal recognition, and global recognition.

**At the individual level,** self-recognition relates to self-confidence and other mindsets related to the individual’s concept of their positionality and capabilities. The pressure to break glass ceilings to achieve success or impact regardless of unfavorable societal and global structures, was expressed. Lack of recognition manifested in compensation structures. Generally, research jobs attract lower pay than clinical jobs which is discouraging for healthcare professionals as the return on investment for research is seemingly low in the short-term. With regard to gender disparities, women acknowledged being less likely to succeed in research given gender roles and multiple demands at the family and societal levels.

“I think it is really overcoming that internalized goliath, we should start early… We have to make people know that you have something to contribute that can make a difference… We have to start that early; we have to change that narrative right from when we are in high school.” [SSA senior male researcher]

“When I come home, I can do some research work or play with my children but for a woman, the same responsibilities plus running the family…, it is not taken very serious by institutions when a woman is asking for research funding, but with a man it is different. It will be a burden on the woman” [SSA senior male researcher]

**At the interpersonal level,** some researchers noted pity or scorn from their counterparts in service delivery. It was disheartening for some to know that, in the sight of their colleagues, they were doomed to failure, but those who maintained their research course found fulfillment in their work.

“When I joined university, it was still very much patriarchal and like very condescending and I don’t see it disappeared because there are still some people who sort of say,” you won’t be able to do that research, it’s impossible.” [SSA senior female researcher]

“My mentor spoke about international collaborations and international publications and that it’s important for your name to be known globally not just to gain money.” [MENA junior female researcher]

**At the institutional level,** institutional recognition was acknowledgment of accomplishments, promotion, and prestige. The lack of recognition of research excellence within institutions undermined the motivation of early career researchers. It was noted that inadequate communication around research outside academia reduces visibility and investment in research which negatively impacts research productivity. The low compensation associated with research roles compared to clinical roles also disincentivized research engagement within institutions. On the other hand, the commercialization of patents was identified as a key benefit of research; however, there was limited researcher capacity to move their work to the marketplace.

“There has to be equity in how much money people are making. For example, if you have two physicians, one is working clinically and one is working as a researcher, the one who is working clinically has double the income of the one who is doing research. They would say, ‘well, what’s the point?’” [SSA senior male researcher]

“The equity goes back also to the value. Do we value research? It goes back to the point that I started with, do we value research, do we value the production of new knowledge.” [MENA senior female researcher]

“Through commercialization… if you have a patent, if you are able to create a company out of your research. That’s another hurdle in the low to middle income countries – the laws, the availability of the infrastructure, even the availability of mentorship in how you move from the lab to the clinical space to the market. Those are skills that very few people have.” [MENA senior female researcher]

**At the national level,** a few participants noted that there is a general preference for a conventional research approaches—mainly dictated by HICs—to doing research which may contribute to the stifling of innovation among early career researchers. One participant pointed out that research has poor public relations (PR) indicating that communication via social media and marketing outlets about the benefits of research has been subpar. Consequently, compared to other institutional mandates, research is not prioritized, and budgetary allocations to run studies, protected research time, or train students are kept to a minimum despite health care being an important sector of society. Beyond institutions, research is not routinely disseminated to lay people and thus its societal relevance is low:

“Let me say we have the western approach to how everything should be done. We are still very focused on certain ways of doing things and disseminating information…What are the other ways in which research can be done, can be disseminated, can be utilized?” [SSA senior male researcher]

“Research has no PR, there is no connect to people either on an individual level or community level or even on a political level and so there is no money for it… you need money for research… There is the money side of research, and the communication side of research.” [SSA senior female researcher]

“There is no connect to people … even on a political level and so there is no money for it… people running research, but they don’t have money. You need money for research and so unfortunately until governments start funding this work, it hard.” [SSA senior female researcher]

**At the global level,** this theme revolved around the lack of global recognition of local researchers and their research (epistemic injustice), power imbalance, and inequities experienced by LMIC researchers (gender, regional, racial, implicit bias, funder bias, and publisher biases). One aspect of global power imbalance, ‘parachute’ science, was identified as a source of inequities in research contributions. The ill effects of parachute research—where foreign researchers come to LMIC or MENA countries to collect data and then leave these countries to write scientific papers using the data without acknowledging the contributions of local researcher—were discussed. This is exemplified in foreign researchers taking leading roles in proposing, implementing and publishing research without incorporating the lived experience and knowledge of local researchers.

“The Western colleagues, I know they want to help us, they want to support us, maybe they have a savior syndrome whatever you call it. You call it they have colonial minds, whatever you call they are trying to help, let us put it that way. They can’t help me because they don’t have the lived-in experience, I am the person living in this experience. I must find my solution, unless they spend time here, they’ll just come in, helicopter …” [ASIA senior male researcher]

“For many of the international collaborations that happened in our region, it was more that they needed partners locally… It involves some capacity building but at the end of the day, really the research is American research or European research… I won’t say that it will necessarily build sufficient capacity. I prefer honestly the more indigenous research—led by people here on the ground.” [MENA senior male researcher]

With regards to global publishing, participants identified inequities in research and bias. Some authors felt that editors were unjustly filtering out manuscript submissions from certain countries especially those from non-English speaking backgrounds. This was cited as a common reason why very few researchers in the developing world have first-author publications or publish in high impact international journals. It also contributes heavily to epistemic injustice—the devaluing of knowledge generated or presented by local researchers. The prohibitive paywalls associated with high-ranking journals often limited their ability to disseminate their research to a wider audience. Most participants noted that without publication fee waivers or collaborative agreements with other researchers from Western institutions, they were unable to afford reputable journals. Other facets of inequities were gender, racial, and career-stage oriented.

“Journals reject and are suspicious of African articles. If I just send an article to the Lancet from Ethiopia and say your name. They will say who is “so and so”? It’s very difficult, but if it’s someone like “John Miller” sends an article from Harvard, I don’t think it will have the same reception.” [SSA senior male researcher]

“In the university that I come from, you have to pay a large portion of the publication fee, especially if it is a high-ranking international journal, out of your own research costs. The university will only cover a portion of it…” [SSA senior female researcher]

“Every time he gets an article reviewed, there’s always a suggestion that it needs to be sent to somebody who’s a native English language. We have our librarians review all of our publications and they’re all native English speakers. There are biases. There are many times you see desk rejections… the same concept applies to having your posters or abstracts accepted at big conferences, or being invited to be a speaker.” [MENA senior female researcher]

## OVERARCHING AND EMERGING THEMES

### 6. Collaboration

Collaboration serves as the catalyst that brings together time, interest, infrastructure, money, expertise and experience, and recognition, to create a thriving research culture even during times of instability (Figure 1). Local and global partnerships, and networks are crucial for developing and supporting a positive research culture.

**Figure 1:**
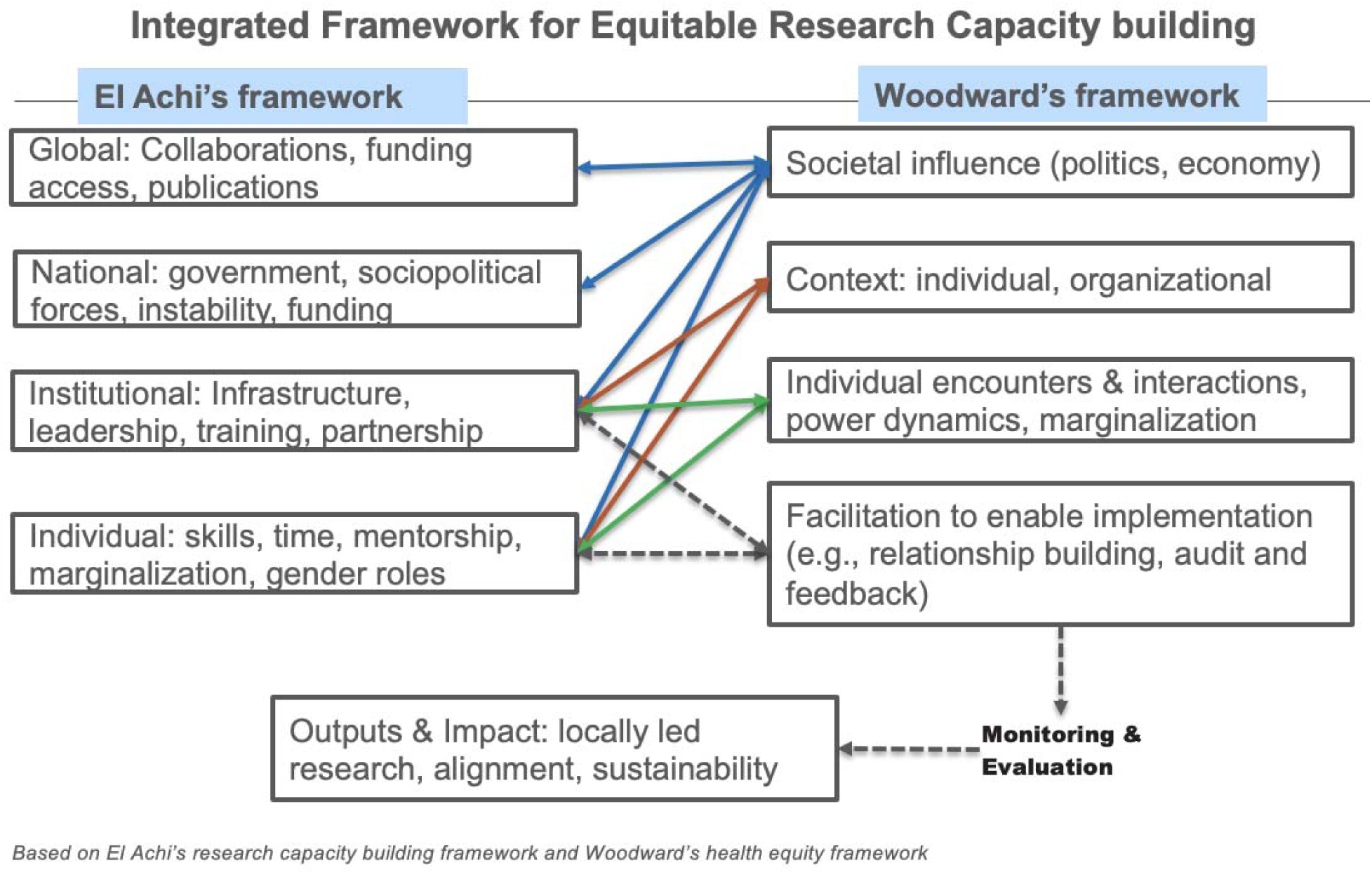
Diagram of integrated conceptual framework.

**At the interpersonal level,** opportunities for collaboration with between emerging and experienced researchers enhance skills and sustain research engagement through mentorship and peer groups. The lack of supportive research communities and circles was something that most researchers had to contend with. Researchers who were well-trained or well-funded found it challenging to integrate into systems where there was no safe structure or circle to lean into and grow. For some, this was one of their most pressing impediments.

“I was able to publish… I got this chance I from Facebook you know. Someone wrote on our Facebook group, “I would like someone interested to help me with research and he will be an author…”There is a research group on Facebook… This helped me a lot because the only way to do research is via connection, Facebook helped me a lot.” [MENA junior male researcher]

“It didn’t have a circle that would allow me to flourish… So why did I lose? I didn’t have the circle that was strong that would allow me to continue to speak about my worth and dream.” [SSA senior female researcher]

**At the institutional and national level,** policies and structures that promote interdisciplinary collaboration strengthen the research culture and expand access to diverse expertise within and across institutions. Participants also stated that national research agendas that encourage pool of resources and expertise can promote collaboration. One participant emphasized the potential of leveraging technology and artificial intelligence (AI) to facilitate collaboration and remote access to resources and networks. Further, participants stated that while collaboration was crucial, the benefits and prestige were largely apportioned to HICs while LMICs bear the load of strenuous field work, with limited or no contribution to the science and publication content.

“Ideas of the projects were coming from high income countries… in low-income countries we just become data collectors, and we would not be mentioned in articles… We put a standing rule that no research will be published without having a local contemporary as a co-author.” [SSA senior male researcher]

“The intention or willingness to collaborate. That’s also quite challenging in the Middle East. I think that’s partly cultural, partly structural as well.” [MENA senior female researcher]

**At the global level,** international collaborations expand research impact through exchange of knowledge, innovations, cross-center trainings, and projects funded for multi-center or multi-country research. Participants emphasized the importance of cultivating equitable research partnerships and forging relationships with funders to be at the cutting edge of emerging opportunities. At the same time, obstacles such as visa and travel restrictions hinder collaboration, making virtual connections and in-country capacity building essential.

“The good thing is now I see in Egypt and Saudia Arabia, I know that they have collaborations with international publishers such as Nature and so on to improve the quality of the research, help the researchers to understand the criteria to publish in such journals. This is helping researchers to improve the quality of their research.” [MENA senior female researcher]

“It’s also about finding the right collaborators, building those personal relationships with the funding agents… what makes you smart is building those relationships with NIH grant program officers, the Gates foundation with the EU, the Canadians, those interpersonal relationships and networks… that’s where you hear information and things you have not seen published online.” [SSA senior male researcher]

“They learn new ways of doing research. They learn about new tools. They learn of ways to overcome the hurdles that they face. This is very important. Not necessarily that they need to go outside the country to do this. Because now there are many challenges facing researchers to go and have their master’s, getting their visa, getting the funding…” [MENA senior female researcher]

### 7. Research Culture

Participants directly or indirectly alluded to *research culture* as either the defining strength of highly productive institutions or the critical gap in low-output settings across SSA and MENA. The prevailing research culture shapes how research is valued, supported, and sustained. In many institutions in these regions, the importance of research, and the benefits it can offer both institutions and communities, is yet to be fully realized. Greater emphasis is needed on the role of research within clinical and teaching settings, particularly in highlighting its short- and longterm benefits for patients, students and institutions.

“At individual level, you are existing in a culture that has dichotomized research as for those with certain degrees versus not. It does not have a system where you can be paid for your time like clinical time.” [SSA senior male researcher]

“I think I would put some of that money in changing behaviors and the mentorship model of potential young researchers. I am not giving up on the old ones but I think I would go to the ones that are still coming up. I will put significant resources in engaging them early…” [SSA senior female researcher]

“Most of the countries in the Middle East are consumers of research and not producers of research. That whole understanding of leading and knowledge, creation of knowledge, is not there. That has several, of course, political, historical, colonial reasons basically behind it.”

[MENA senior female researcher]

“As part of this building, the ecosystem or the scientific community, they even give funding for high school students to do research and opportunities for internships. They actually ask us to go to schools sometimes to talk about research.” [MENA senior male researcher]

“This is a common understanding that research is not needed as a physician… it is a common culture. This is something that you are just doing for grades, to pass that year… we don’t see the value of it, we don’t use it, we don’t apply it.” [MENA junior male researcher]

“…the culture of the place is so you cannot do any research that makes the government look bad. There are a lot of health issues that are really bad, but why would I research and pose problems?” [MENA senior male researcher]

## MODIFIED DELPHI APPROACH RANKING AND RESULTS

Top factors exposed through the modified Delphi process are outlined and displayed in the heat map (Table 1) and mapped onto the *TIMER-2C framework*. Colors were assigned to each barrier from 1-5 based on the number of votes received. Deeper green shades indicate a higher number of votes for a specific ranking (higher importance), while orange shades represent fewer votes, and red shades indicate the least number of votes.

### At the individual level

Key barriers: Researchers ranked expertise and experience gaps (such as lack of opportunities, lack of training, and language barriers), lack of passion, confidence and commitment, and insufficient funding as the most significant hurdles/barriers.

Key facilitators identified were passion and commitment, confidence, protected time, and training to promote research competency.

### At the interpersonal level

<u>Key barriers identified were</u> limited funding in competitive environments, inequitable recognition (patriarchy, gender bias, hierarchy, and power imbalances), and scarcity of protected research time. <u>Key facilitators included</u> training and mentorship.

### At the institutional level

<u>Key barriers were</u> lack of dedicated research funding and limited protected time which stems from the lack of a research culture. <u>Key facilitators were</u> building a research culture and sustainable research pipeline in addition to creating supportive institutional policies (around promotion, compensation and incentives, research administration, and funding).

### At the national or wider ecosystem level

Key barriers identified were limited government funding and commitment, lack of research importance, poor communication and public relations, while key facilitators were supporting research dissemination nationally to increase its visibility and recognition, and establishing national mechanisms for research funding, and fostering collaboration across institutions.

### At the global level

Bias, inequity in publishing, funding, epistemic injustice, and parachute science were key barriers, and key facilitators were equitable collaborations and partnerships, scholarship and exchange programs, and autonomous funding free of external funder control and agendas.

**APPENDIX 2:** outlines the key themes, codes, and sample quotes.

The overarching theme of “research culture,” was mapped onto six key domains or themes for the key determinants of research productivity: **T**ime, **I**nstability, Interest and Infrastructure, **M**eans of funding research, **E**xpertise & **E**xperience, **R**ecognition, and **C**ollaboration, which we summarized by the mnemonic **TIMER-2C** hence the *“TIMER-2C Conceptual Framework for Research Culture.”* (Figure 1)

**Figure 1:**
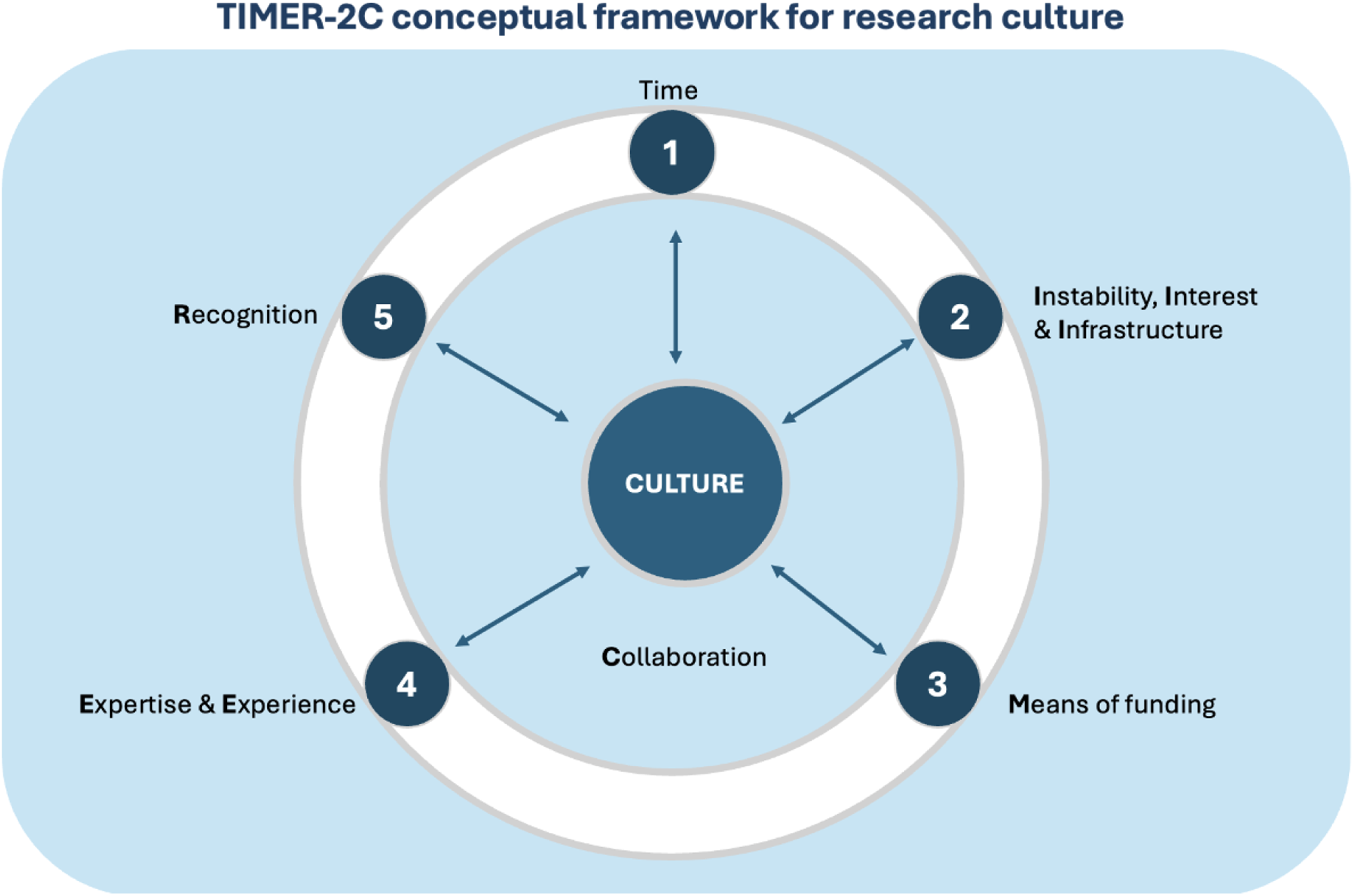
TIMER- 2C Conceptual Framework for Research Culture.

## DISCUSSION

We sought to understand determinants of local research productivity in the MENA and SSA, and broadly in low income countries (LICs), and middle income countries (MICs). Based on our findings, we unveil an emerging conceptual framework of determinants, the *TIMER-2C framework for Research Culture*, that explains the relationship between themes associated with research productivity—specifically: 1) time; 2) instability, interest and infrastructure 3) means of funding research; 4) expertise and experience; 5) recognition; 6) constructive collaboration; and 7) culture. In our synthesis, research culture emerged as either the defining strength of highly productive healthcare professionals or the critical gap hindering research output across SSA and MENA. Research culture cuts across all levels—from individual to national—and shapes how research is valued, supported, and sustained. In many settings, research is seen as academically perfunctory or irrelevant to daily professional roles and is given less priority than mainstream clinical service delivery and teaching. Overall, we found that research culture is shaped by competing priorities including time, instability, interest, infrastructure, funding, and a lack of expertise, experience, recognition, and collaboration.

We found that experiences such as joining collaborative projects were vital to gain research skills, funding, mentorship, publications and recognition. However, dissatisfaction with inequitable partnerships that disenfranchise local researchers was evident.^19^ This is more concerning among women who are underrepresented in research participation.^6,9^ A great opportunity exists to reshape research culture by addressing these factors and positioning health research as *a societally recognized cornerstone for healthcare, academia, and public health.* While there have been regional and international initiatives to improve research productivity in LMICs, these efforts have been variable, targetting various aspects of research capacity building with few addressing the multi-level dimensions of research culture and the underlying structural inequities that undermine research productivity.

**There is an urgent need for frameworks to evaluate, assess and promote equitable research culture** in low output settings like the MENA and SSA. To our knowledge, there are no frameworks—that are grounded in lived experiences and recommendations from local researchers—that evaluate the robustness of research culture and its influences at the multiple socioecological levels. We posit that the barriers and facilitators of a positive research culture are multi-level encompassing individual, interpersonal, institutional, government, and global dimensions. As such, multi-level approaches and strategies are needed to address the complex web of factors affecting research culture and productivity.^20^ Understanding how these factors map onto the key themes of the *TIMER-2C framework* will aid in crafting relevant interventions. In this light, our synthesis of the TIMER-2C framework, serves as a foundational step for rigorous evaluation and tailoring of informed interventions to improve research contributions from local healthcare professionals in the MENA, SSA and other LMICs.

Existing health research capacity strengthening (RCS) **frameworks have focused on research capacity building, often overlooking the *culture of research* in LMICs** and few look into the perspectives from marginalized groups and unstable settings.^19,21^ These frameworks are complex, often driven by funder requirements, and lack emphasis on contextual factors. A study of seven frameworks from 2004 to 2012 found that they were primarily used for internal organizational performance and made limited reference to specific theories.^22^ The study suggested that frameworks could be improved by incorporating practical guidance, engaging more stakeholders, and better harmonization of concurrent efforts. The TIMER-2C framework builds on these deficiencies to ensure that practical and contextual perspectives from local stakeholders are represented and provides a clear agenda for areas of prioritization.

**Some frameworks have primarily been guided by the priorities of funders and external institutions**. The ESSENCE Research Capacity Building framework^23^ was developed in 2014– 2016—through a consultative process led by the ESSENCE on Health Research initiative, an initiative by funders and bilateral development agencies to synergize, establish coherence, and coordinate resources for health research in LMICs^24,25^—to harmonize stakeholders in alignment with health priorities and sustain research strengthening beyond individual skills. ESSENCE is a more prescriptive, investment-oriented framework, whereas TIMER-2C is diagnostic, context-specific framework centered on the end-user or action target—the researcher. TIMER-2C’s nuance comes from grounding in the lived experiences of local healthcare professionals navigating local and global research structures, thus can greatly complement the approaches – and counter the limitations – of existing frameworks and previous top-down research capacity building efforts.

Cooke et al. used **a realist synthesis approach to analyze literature** – primarily from HIC settings – on research capacity strengthening from 36 conceptual and theoretical papers to identify their underlying mechanisms.^26^ Similar to the TIMER-2C framework, the realist synthesis identified eight program theories that distill mechanisms operating at multiple levels (individual, team, and organizational). Mechanisms were classified as either symbolic (i.e., fulfill a symbolic role by elevating the importance of research capacity development e.g., signaling importance, positive role models) or functional (i.e., practical mechanisms designed to actively build research skills and culture e.g., learning by doing). Out of 36 studies, only six were relevant to our context—four with LMIC focus, one from Liberia, and one from South Africa. While Cooke et al. offers a mechanistic lens on how change occurs, the TIMER-2C framework complements this by diagnosing what needs to change and where—at which socio-ecological levels—those changes should be targeted.

Overarchingly, **aspects of power asymmetries and instability in RCS are less accounted for hence the need for an integrated framework like TIMER-2C.** One study found that progress was hindered by broader structural inequalities in an RCS program, illustrating that focus on individual research careers was not sufficient.^19^ TIMER-2C frames this more explicitly under the “Recognition” theme which situates the problem across multiple levels—not only the individual and institutional levels, but also the national and global mechanisms (authorship norms, publication, funding flows, agenda-setting) that perpetuate inequities. We demonstrate how this manifests, providing a basis for potential mitigation strategies. El Achi et al. provides a conceptual framework for RCS, addressing structural levels and highlights conflict-specific constraints—safety, political permissibility, disrupted infrastructure and communications, and data restrictions—as determinants of success.^16^ We draw on this framework’s emphasis on multi-level dynamics and external sociopolitical context, and expand beyond this to the global ecosystem.

**There are varied efforts to promote research culture in LMICs.**^27^ We found variability in supportive institutional cultures across countries in MENA and SSA. A positive research culture comprises the environment, values, norms, and practices that shape the behavior and attitudes within healthcare, academia or research institutions, including the prevailing tendencies and standards that guide how research is conducted, communicated, and evaluated.^28^ We provide three recommendations that target factors affecting multiple levels of research culture and productivity: 1) Establishing learning communities and safe research circles with opportunities for research capacity building (this targets individual/ interpersonal and institutional levels); 2) autonomous leadership and funding untethered to politics or external forces (this is relevant to institutional, national levels, and global levels); and 3) creating sustainable pipelines of early career researchers (individual/ interpersonal, institutional and global levels). The most significant barriers were posed by institutional structures (the boiling pot in which many challenges converged), thus our multi-level recommendations target the institutional level in conjunction with one or more additional levels – higher or lower.

**Establishing learning communities and safe research circles with equitable opportunities** for research capacity building should be the goal for institutions seeking to nurture and promote research. Passion and interest in research are only as good as the equitable opportunities—for research, training, mentorship, career, and capacity building—afforded to someone.^29^ For example, Edwards et al. found that when researchers are given opportunities to work alongside senior researchers, in-person or remotely, they gain skills to balance research with teaching, clinical and administrative demands.^30,13^ Successful programs like the Fogarty Global Health Fellows program funded by the National Institute of Health (NIH), the Medical Education Partnership Initiative (MEPI) and the CARTA program for African scientists serve as a blueprint for establishing research communities and hubs that connect trainees and mentees to clear career trajectories.^27,31,32,33^ However, conventional training programs do not necessarily cater for post-training career pathways or funding resulting in junior researcher isolation, siloing or exiting the field. More thought is needed on where to transplant and nurture junior researchers alongside established researchers. This is aggravated by brain drain, a vicious cycle triggered by lack of opportunities and perpetuated by the loss of skilled professionals. As individuals seek training abroad and opt to remain overseas, their absence further weakens the home country’s capacity to develop and retain experts. To curb brain drain, there should be a focus on nurturing junior researchers’ and providing attractive career opportunities with clear growth trajectories.^30^ In our synthesis, we established that stepwise approaches to optimize the immediate research environment of early career or junior researchers (the individual level) and sequentially expand to higher levels at the institution and broader ecosystem can be most feasible.

**We posit that autonomous funding models and leadership,** free from external political or governmental agendas, can significantly transform the research culture. We recognize that while autonomous funding is not a straightforward solution, there has been progress in this regard.^34,35,36^ We found that the biggest obstacles emanate from the institutional level, where leadership priorities and funding availability dictate access to protected time, and other pre-requisites for successful research.^37^ Our data revealed that thriving learning research communities escaped the “funding trap”—complete reliance on donors, institutional funds and administrative structures—through autonomous funding mechanisms and low-cost innovation,^34^ while maintaining close partnerships with parent institutions for infrastructure support.^38^ One recommendation is to establish local research funds managed by independent boards with contributions – e.g., from diaspora philanthropists, local business through corporate social responsibility arms, and local governments and partnerships between regional universities– could ensure stability despite political cycles.^36,39,40^

**Building a sustainable pipeline of researchers** is another important strategy grounded in all the determinants outlined in the TIMER-2C framework. Research needs to be introduced early in formal education, particularly among women. Partnership programs that extend to high schools or colleges are needed to establish continuous entry points for trainees into research. Using “Trainer of Trainee” models can greatly boost the sustainability of research by preparing the stage for the next generation of research leaders.^41^

**The question of how to leverage Artificial Intelligence (AI) and technology** to surmount these gaps arises especially with regard to *establishing learning communities* and *sustainable pipeline of researchers*. For example:

a. Administrative simplification: AI has a huge role to play in making the research field even for MENA and SSA researchers by automating routine administrative and supportive tasks and thus free up time for higher-level tasks to do with scientific inquiry, innovation, and conceptualization of research. Furthermore, AI can support administrative tasks such as budget generation, grant writing, reducing the administrative burden on researchers.
b. Research enhancement: AI tools can also streamline data collection (e.g., adapting data collection tools), data analysis and validation.
c. Infrastructure and collaboration: AI and technological advances have the potential to mitigate the impact of instability and infrastructure challenges by enabling remote collaboration and data sharing.
d. Quality improvement: Other roles include improving the quality of research by assisting with language editing and proofreading.

While these facilitators can promote interest and engagement in research by healthcare providers, we recognize that majority of these benefits depend on reliable internet broadband access which may not be guaranteed in fragile regions.

**The TIMER-2C framework could be leveraged as a practical tool for monitoring and evaluation of positive research culture** within RCS programs by standardizing indicators for each theme. For example, *Time* could be assessed by the proportion of staff with protected research time; *Recognition* by the presence, number, and type of institutional incentives and awards; and *Culture* by staff perceptions of research value and the number of policy or practice changes resulting from research initiatives. These indicators could be combined into an index to evaluate institutional research culture, supporting both baseline assessments and ongoing tracking of progress over time.

**Our work is not without limitations.** First, we experienced a relatively high non-response rate (∼50%), which may have limited the full extent of the breadth of our responses. The limited number of participants inevitably constrains generalizability. Our purposive sampling strategy was intentionally designed to prioritize depth of experiences over representativeness. Second, our predominantly qualitative approach restricted the robustness and validation of the findings. Third, we acknowledge that the perspectives captured may not fully represent the diversity of SSA research contexts, particularly in non–English-speaking regions like Central Africa.

**Key strengths** of our study include the diverse representation of participants from multiple countries across both regions, providing a diverse range of perspectives and equal contributions by sex. Second, we were able to capture insights from both emerging and established researchers. Third, the qualitative approach, guided by integrated theoretical frameworks, offered a holistic lens for exploring perspectives, and transcripts were returned to participants for review and verification to enhance data accuracy. Fourth, we compared this framework with other conceptual frameworks to understand the gaps, and relevance, of the TIMER-2C framework in addressing these gaps. Fifth, the modified Delphi method, was valuable for building consensus and enabled quantitative ranking of key factors at various levels.

## CONCLUSION

**In conclusion**, strengthening research culture and productivity in MENA and SSA requires a clear, actionable roadmap—and the emerging TIMER-2C framework offers that. The TIMER-2C framework can serve as a multi-level evaluation tool to assess the progress and impact of both existing and future initiatives aimed at enhancing research culture and productivity. Future directions include further testing of the framework on published data from our ongoing systematic review on the same topic, validation, and contextual adaptation to enable individuals, institutions, and countries to implement effective strategies for improving research output and productivity.

## Data Availability

All data produced in the present study are available upon reasonable request to the authors

## Acknowledgements

We would like to acknowledge all the key informants who agreed to participate in these interviews and modified Delphi process. We also acknowledge the Washington University research team who helped with the administrative processes needed to complete this work.

## Author contributions

Beryne Odeny is responsible for the overall content as guarantor.

Beryne Odeny: conception, design, data collection, analysis, interpretation of the data, drafting the work and revision of the manuscript.

Dorothy Mangale: data collection, analysis, interpretation of the data, drafting the work and revision of manuscript.

Dhananjaya Sharma: interpretation of data, and review of manuscript Mariam Balogun: interpretation of data and drafting of the work Isaac Che Ngang: interpretation of the data and review of manuscript

Vaishnavi Mamillapalle: interpretation of data, and drafting of manuscript Akila Anandarajah: interpretation of data, revision and writing of manuscript Ermyas Biiru: interpretation of data and review of the manuscript

Fatima Suleman: interpretation of data and review of the manuscript Miliard Derbew: interpretation of data and review of the manuscript Juliet Iwelunmor: interpretation of data and review of the manuscript

Wael Al Mahmeed: acquisition, conception, interpretation of data, and review of manuscript.

All authors critically reviewed the manuscript and gave final approval of the manuscript to be published.

## Abbreviations

AI: Artificial Intelligence
COREQ: Consolidated Criteria for Reporting Qualitative Research
HICs: High-Income Countries
LMICs: Low- and Middle-Income Countries
MENA: Middle East and North Africa
NIH: National Institutes of Health
PR: Public Relations
SSA: sub-Saharan Africa
TIMER-2C: Time, Instability/Interest/Infrastructure, Means of funding, Expertise/Experience, Recognition, Collaboration and Culture

## Appendix 1: COREQ checklist (attached as a supplementary file)

## Appendix 2: SAMPLE CODES AND EXEMPLARY QUOTES BY THEME

**Table 1:**
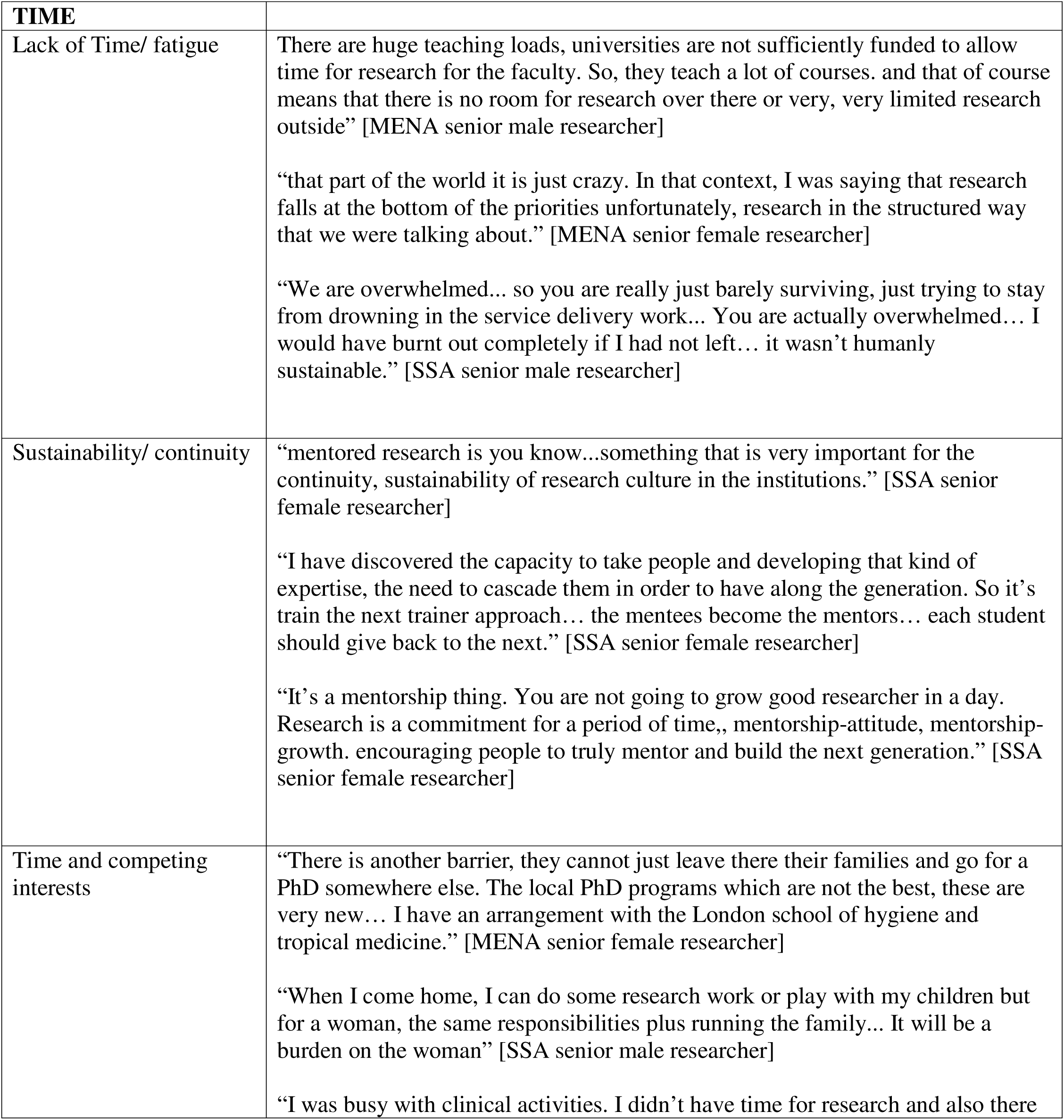
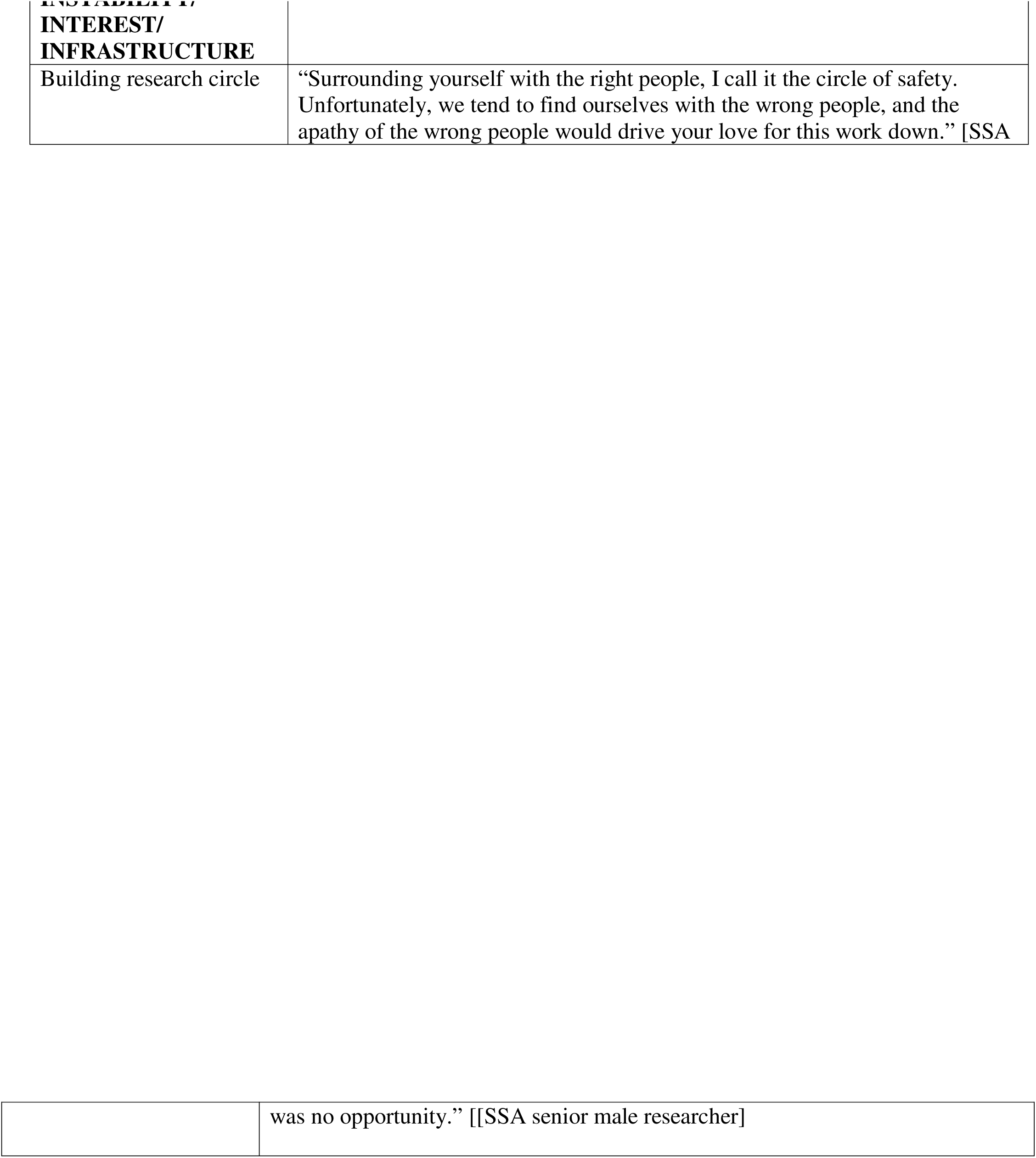
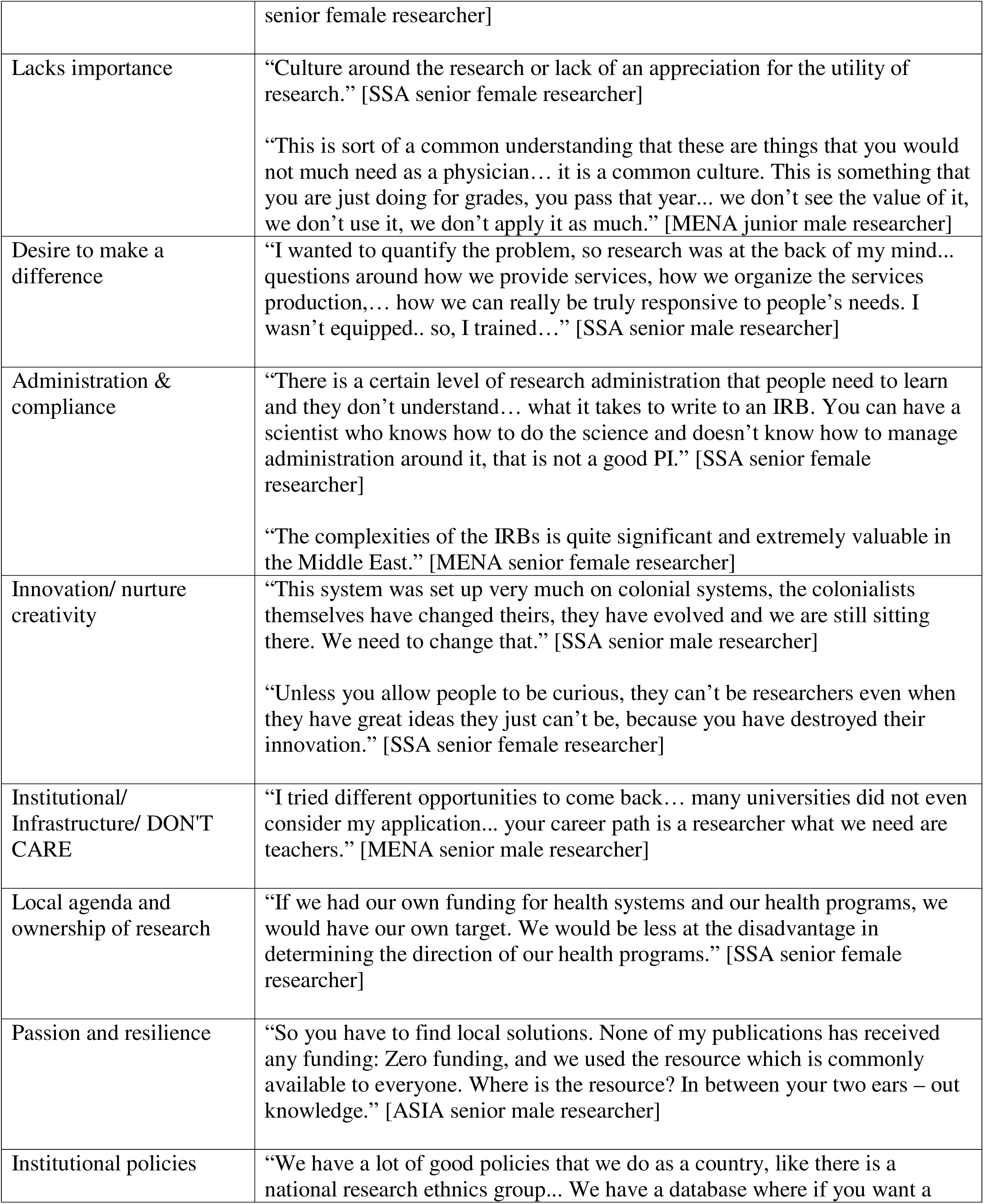
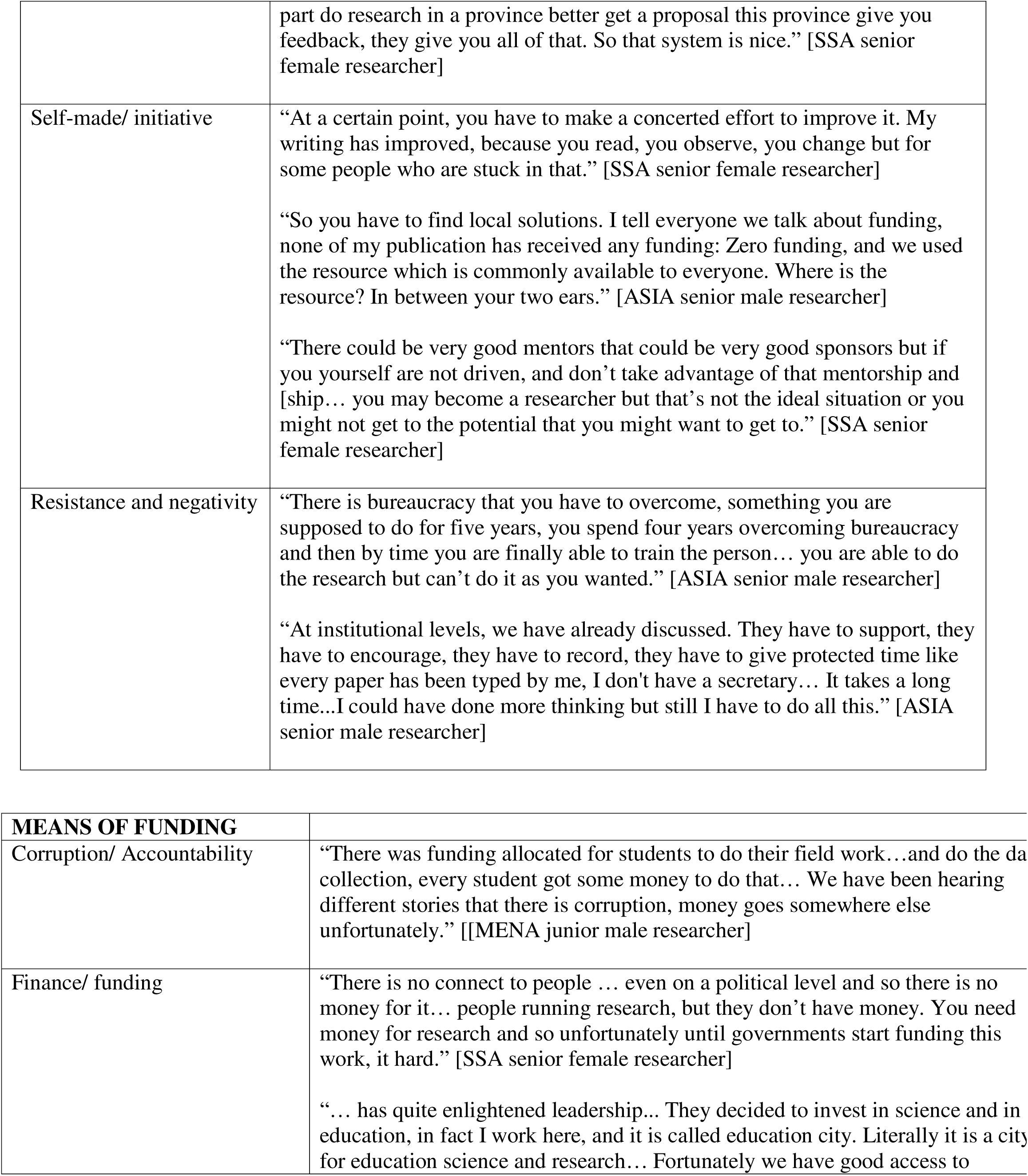
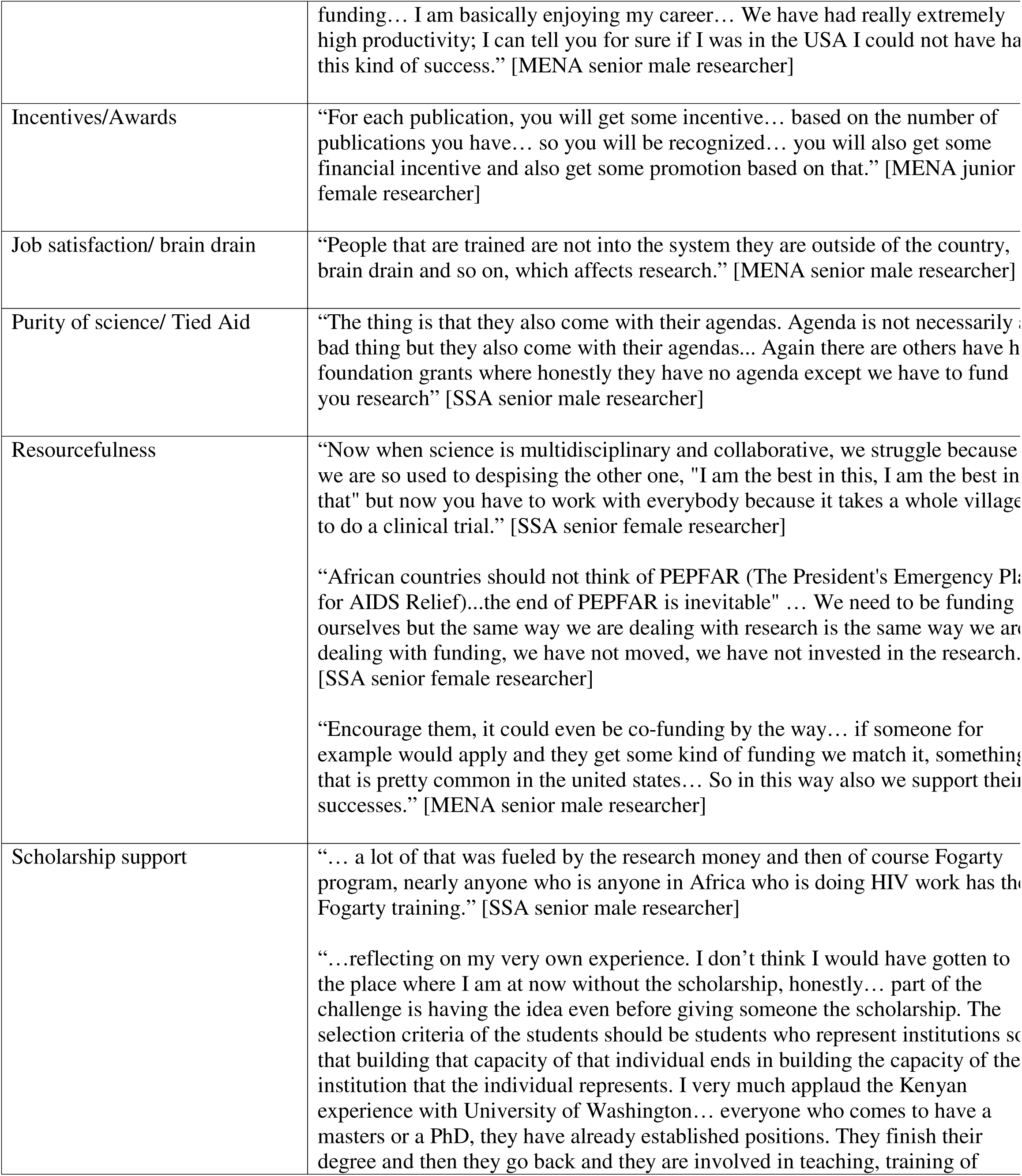
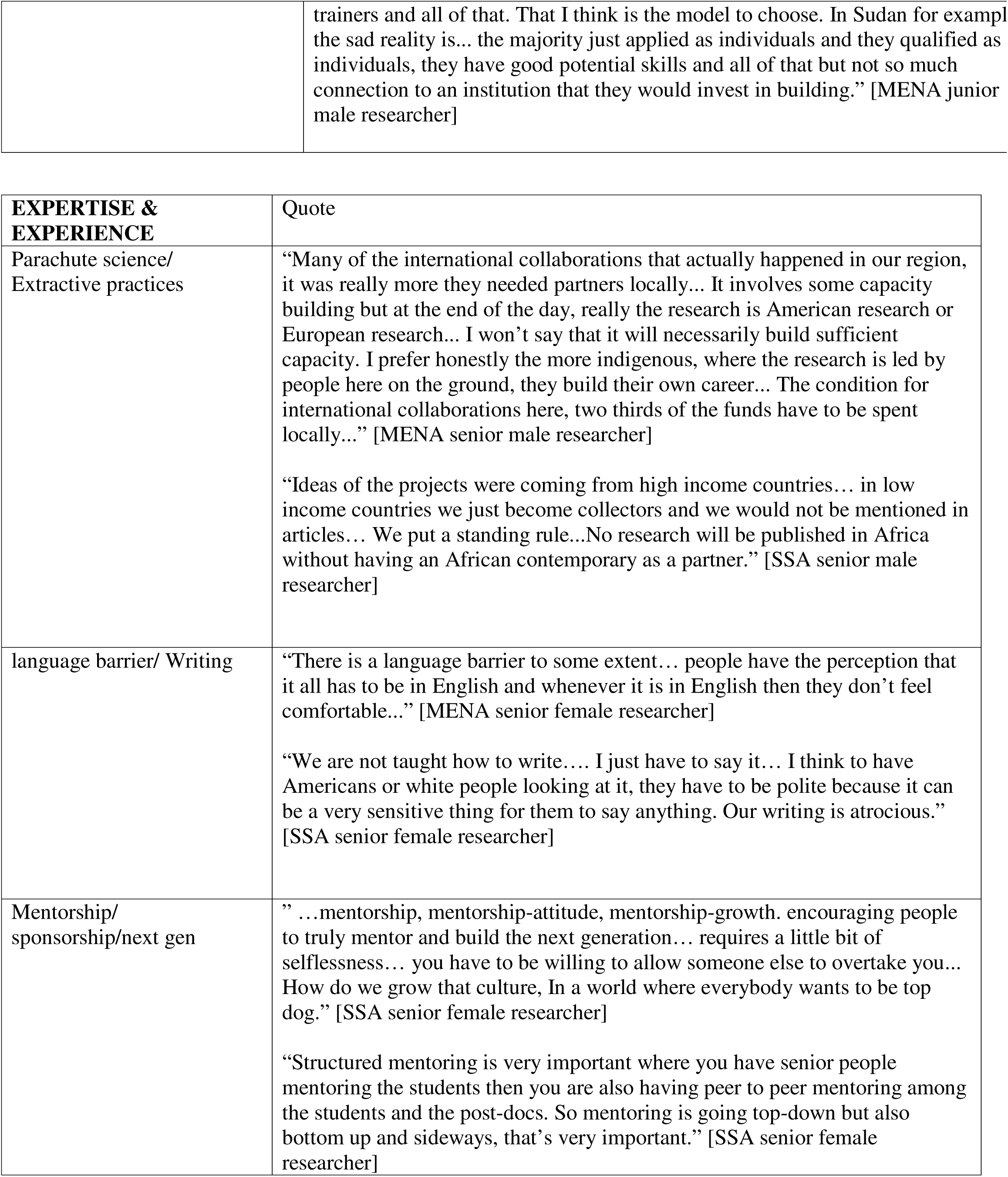
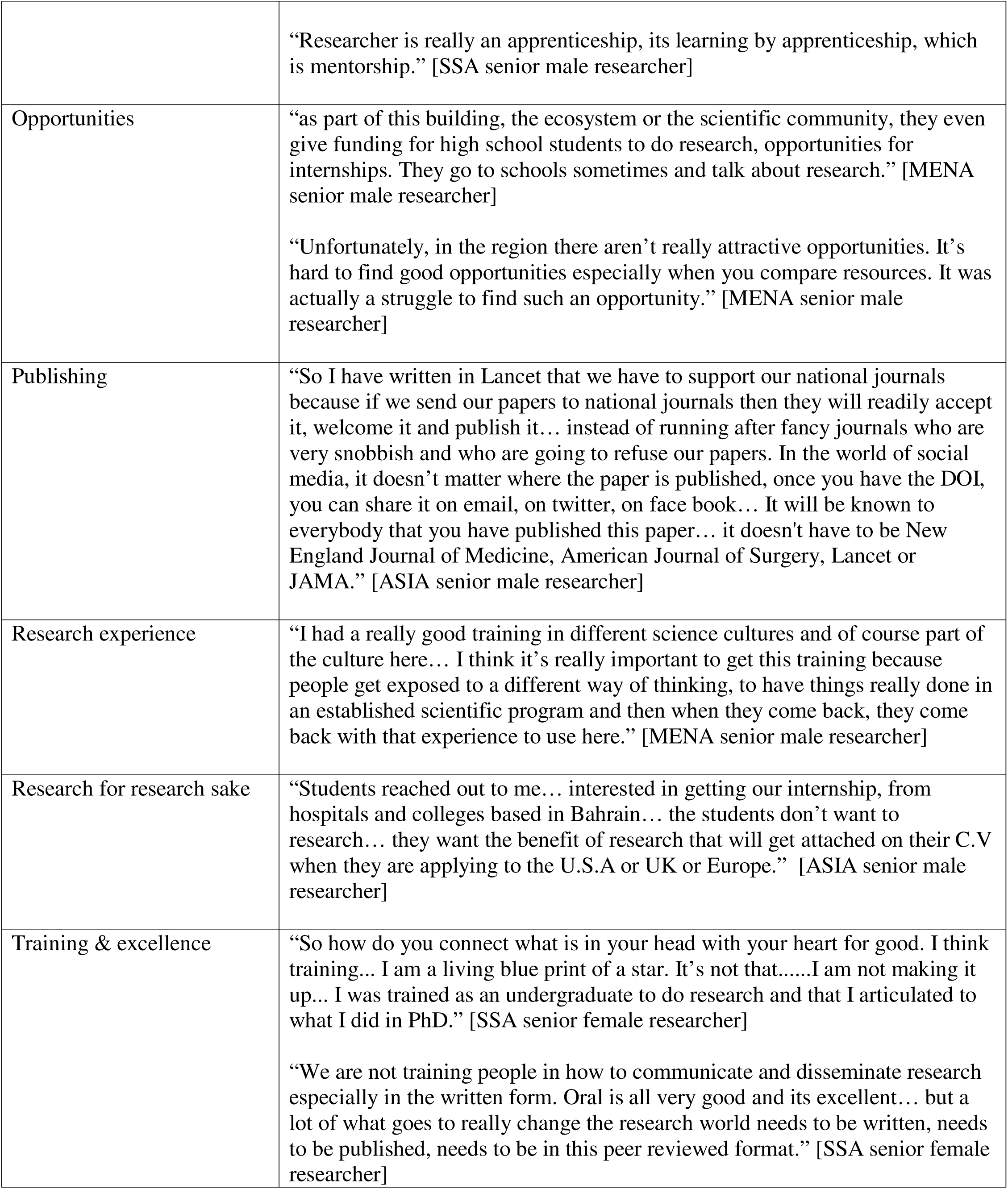
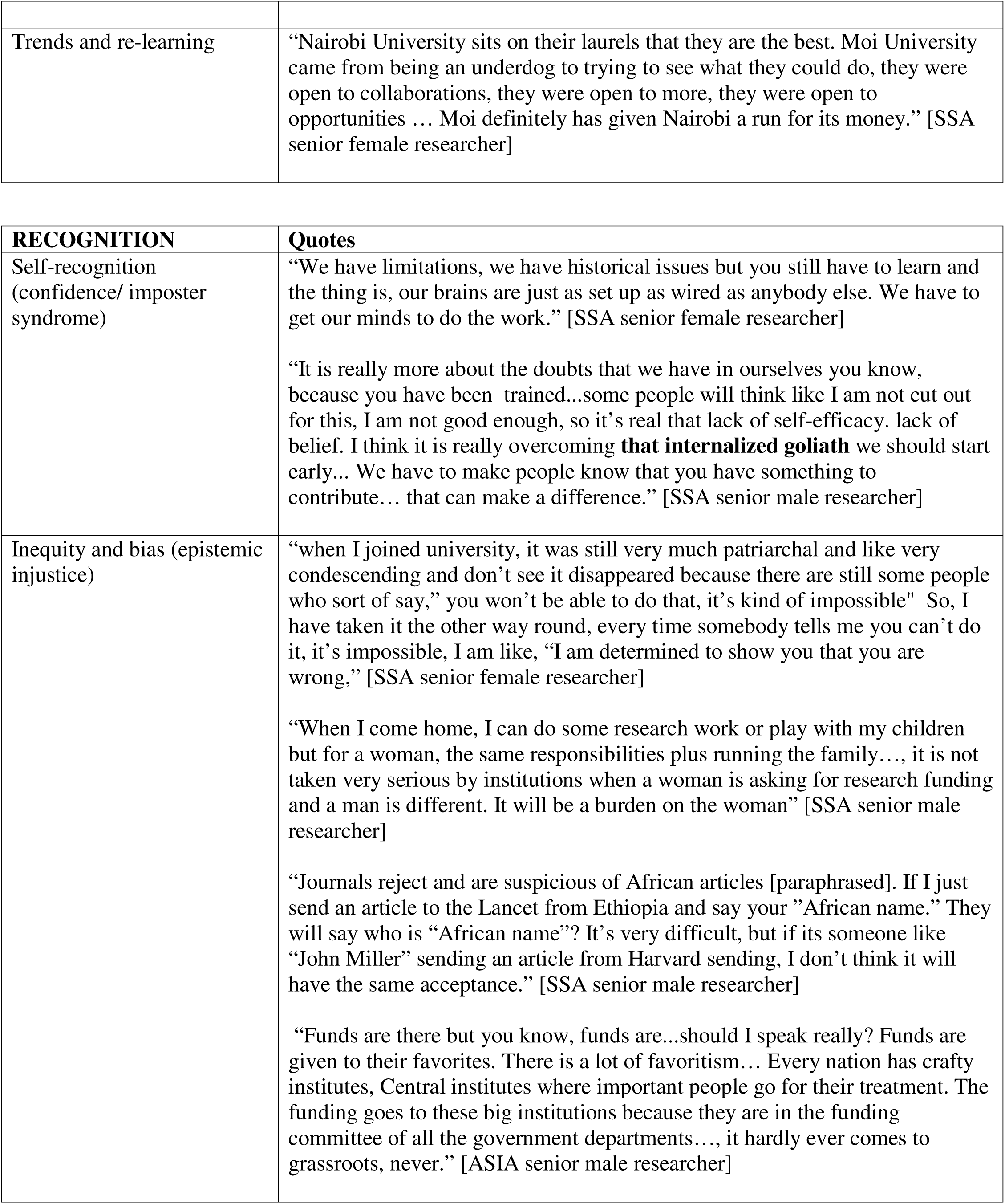
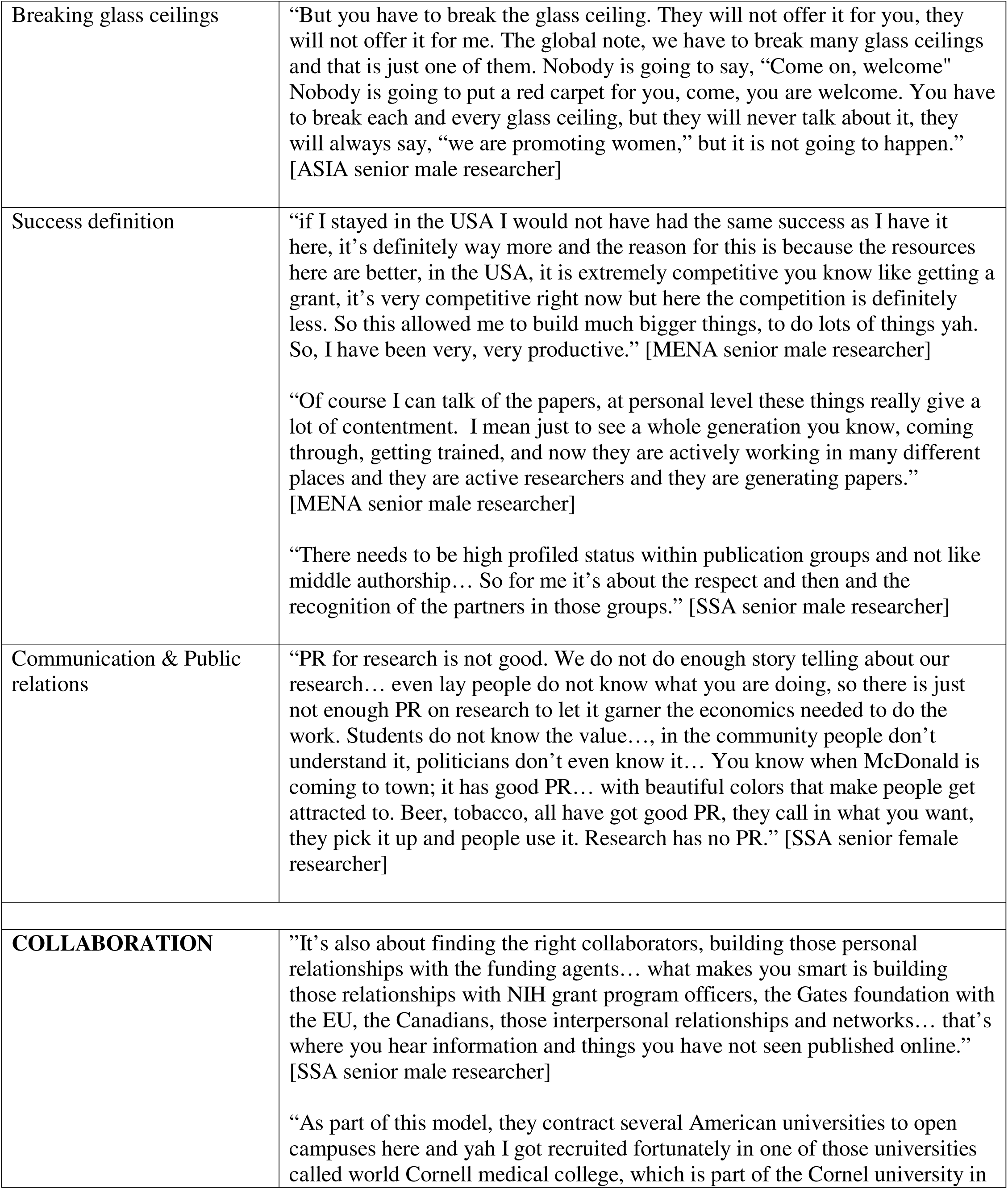
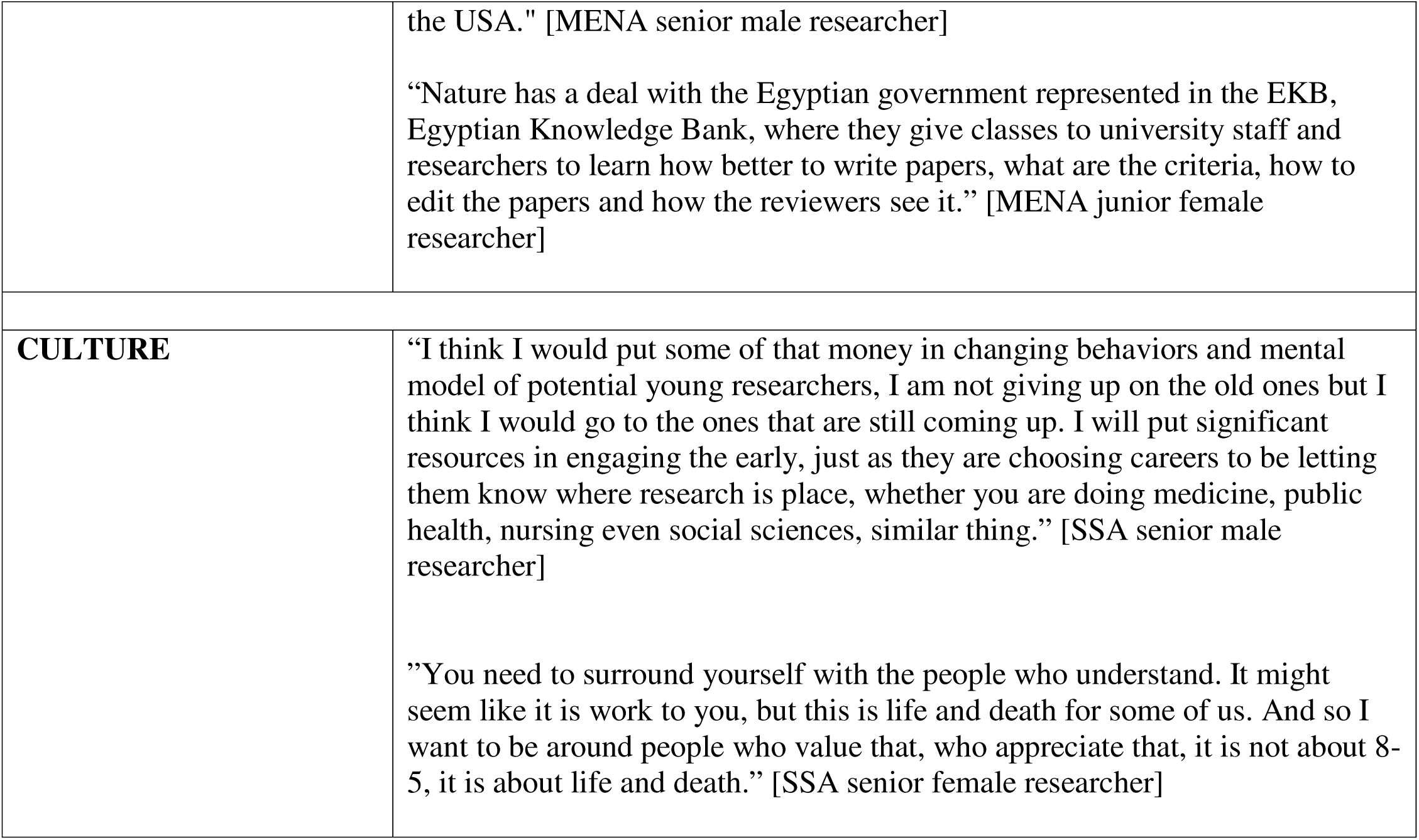
Participant characteristics.

| TIME |  |
| --- | --- |
| Lack of Time/ fatigue | There are huge teaching loads, universities are not sufficiently funded to allow time for research for the faculty. So, they teach a lot of courses. and that of course means that there is no room for research over there or very, very limited research outside” [MENA senior male researcher]<br>“that part of the world it is just crazy. In that context, I was saying that research falls at the bottom of the priorities unfortunately, research in the structured way that we were talking about.” [MENA senior female researcher]<br>“We are overwhelmed... so you are really just barely surviving, just trying to stay from drowning in the service delivery work... You are actually overwhelmed... I would have burnt out completely if I had not left... it wasn’t humanly sustainable.” [SSA senior male researcher] |
| Sustainability/ continuity | “mentored research is you know...something that is very important for the continuity, sustainability of research culture in the institutions.” [SSA senior female researcher]<br>“I have discovered the capacity to take people and developing that kind of expertise, the need to cascade them in order to have along the generation. So it’s train the next trainer approach... the mentees become the mentors... each student should give back to the next.” [SSA senior female researcher]<br>“It’s a mentorship thing. You are not going to grow good researcher in a day. Research is a commitment for a period of time,, mentorship-attitude, mentorship-growth. encouraging people to truly mentor and build the next generation.” [SSA senior female researcher] |
| Time and competing interests | “There is another barrier, they cannot just leave there their families and go for a PhD somewhere else. The local PhD programs which are not the best, these are very new... I have an arrangement with the London school of hygiene and tropical medicine.” [MENA senior female researcher]<br>“When I come home, I can do some research work or play with my children but for a woman, the same responsibilities plus running the family... It will be a burden on the woman” [SSA senior male researcher]<br>“I was busy with clinical activities. I didn’t have time for research and also there |
| **INTEREST/**<br>**INFRASTRUCTURE** |  |
| Building research circle | “Surrounding yourself with the right people, I call it the circle of safety. Unfortunately, we tend to find ourselves with the wrong people, and the apathy of the wrong people would drive your love for this work down.” [SSA |
|  | was no opportunity.” [[SSA senior male researcher] |
|  | senior female researcher] |
| Lacks importance | “Culture around the research or lack of an appreciation for the utility of research.” [SSA senior female researcher]<br>“This is sort of a common understanding that these are things that you would not much need as a physician... it is a common culture. This is something that you are just doing for grades, you pass that year... we don’t see the value of it, we don’t use it, we don’t apply it as much.” [MENA junior male researcher] |
| Desire to make a difference | “I wanted to quantify the problem, so research was at the back of my mind... questions around how we provide services, how we organize the services production,... how we can really be truly responsive to people’s needs. I wasn’t equipped.. so, I trained...” [SSA senior male researcher] |
| Administration & compliance | “There is a certain level of research administration that people need to learn and they don’t understand... what it takes to write to an IRB. You can have a scientist who knows how to do the science and doesn’t know how to manage administration around it, that is not a good PI.” [SSA senior female researcher]<br>“The complexities of the IRBs is quite significant and extremely valuable in the Middle East.” [MENA senior female researcher] |
| Innovation/ nurture creativity | “This system was set up very much on colonial systems, the colonialists themselves have changed theirs, they have evolved and we are still sitting there. We need to change that.” [SSA senior male researcher]<br>“Unless you allow people to be curious, they can’t be researchers even when they have great ideas they just can’t be, because you have destroyed their innovation.” [SSA senior female researcher] |
| Institutional/ Infrastructure/ DON'T CARE | “I tried different opportunities to come back... many universities did not even consider my application... your career path is a researcher what we need are teachers.” [MENA senior male researcher] |
| Local agenda and ownership of research | “If we had our own funding for health systems and our health programs, we would have our own target. We would be less at the disadvantage in determining the direction of our health programs.” [SSA senior female researcher] |
| Passion and resilience | “So you have to find local solutions. None of my publications has received any funding: Zero funding, and we used the resource which is commonly available to everyone. Where is the resource? In between your two ears – out knowledge.” [ASIA senior male researcher] |
| Institutional policies | “We have a lot of good policies that we do as a country, like there is a national research ethnics group... We have a database where if you want a |
|  | part do research in a province better get a proposal this province give you feedback, they give you all of that. So that system is nice.” [SSA senior female researcher] |
| Self-made/ initiative | “At a certain point, you have to make a concerted effort to improve it. My writing has improved, because you read, you observe, you change but for some people who are stuck in that.” [SSA senior female researcher]<br>“So you have to find local solutions. I tell everyone we talk about funding, none of my publication has received any funding: Zero funding, and we used the resource which is commonly available to everyone. Where is the resource? In between your two ears.” [ASIA senior male researcher]<br>“There could be very good mentors that could be very good sponsors but if you yourself are not driven, and don’t take advantage of that mentorship and [ship... you may become a researcher but that’s not the ideal situation or you might not get to the potential that you might want to get to.” [SSA senior female researcher] |
| Resistance and negativity | “There is bureaucracy that you have to overcome, something you are supposed to do for five years, you spend four years overcoming bureaucracy and then by time you are finally able to train the person... you are able to do the research but can’t do it as you wanted.” [ASIA senior male researcher]<br>“At institutional levels, we have already discussed. They have to support, they have to encourage, they have to record, they have to give protected time like every paper has been typed by me, I don't have a secretary... It takes a long time...I could have done more thinking but still I have to do all this.” [ASIA senior male researcher] |

| **MEANS OF FUNDING** |  |
| --- | --- |
| Corruption/ Accountability | “There was funding allocated for students to do their field work...and do the data collection, every student got some money to do that... We have been hearing different stories that there is corruption, money goes somewhere else unfortunately.” [[MENA junior male researcher] |
| Finance/ funding | “There is no connect to people ... even on a political level and so there is no money for it... people running research, but they don’t have money. You need money for research and so unfortunately until governments start funding this work, it hard.” [SSA senior female researcher]<br>“... has quite enlightened leadership... They decided to invest in science and in education, in fact I work here, and it is called education city. Literally it is a city for education science and research... Fortunately we have good access to |
|  | funding... I am basically enjoying my career... We have had really extremely high productivity; I can tell you for sure if I was in the USA I could not have had this kind of success.” [MENA senior male researcher] |
| Incentives/Awards | “For each publication, you will get some incentive... based on the number of publications you have... so you will be recognized... you will also get some financial incentive and also get some promotion based on that.” [MENA junior female researcher] |
| Job satisfaction/ brain drain | “People that are trained are not into the system they are outside of the country, brain drain and so on, which affects research.” [MENA senior male researcher] |
| Purity of science/ Tied Aid | “The thing is that they also come with their agendas. Agenda is not necessarily a bad thing but they also come with their agendas... Again there are others have had foundation grants where honestly they have no agenda except we have to fund your research” [SSA senior male researcher] |
| Resourcefulness | “Now when science is multidisciplinary and collaborative, we struggle because we are so used to despising the other one, "I am the best in this, I am the best in that" but now you have to work with everybody because it takes a whole village to do a clinical trial.” [SSA senior female researcher]<br>“African countries should not think of PEPFAR (The President's Emergency Plan for AIDS Relief)...the end of PEPFAR is inevitable" ... We need to be funding ourselves but the same way we are dealing with research is the same way we are dealing with funding, we have not moved, we have not invested in the research. [SSA senior female researcher]<br>“Encourage them, it could even be co-funding by the way... if someone for example would apply and they get some kind of funding we match it, something that is pretty common in the united states... So in this way also we support their successes.” [MENA senior male researcher] |
| Scholarship support | “... a lot of that was fueled by the research money and then of course Fogarty program, nearly anyone who is anyone in Africa who is doing HIV work has the Fogarty training.” [SSA senior male researcher]<br>“...reflecting on my very own experience. I don't think I would have gotten to the place where I am at now without the scholarship, honestly... part of the challenge is having the idea even before giving someone the scholarship. The selection criteria of the students should be students who represent institutions so that building that capacity of that individual ends in building the capacity of the institution that the individual represents. I very much applaud the Kenyan experience with University of Washington... everyone who comes to have a masters or a PhD, they have already established positions. They finish their degree and then they go back and they are involved in teaching, training of |
|  | trainers and all of that. That I think is the model to choose. In Sudan for example the sad reality is... the majority just applied as individuals and they qualified as individuals, they have good potential skills and all of that but not so much connection to an institution that they would invest in building.” [MENA junior male researcher] |

| **EXPERTISE & EXPERIENCE** | **Quote** |
| --- | --- |
| Parachute science/<br>Extractive practices | “Many of the international collaborations that actually happened in our region, it was really more they needed partners locally... It involves some capacity building but at the end of the day, really the research is American research or European research... I won’t say that it will necessarily build sufficient capacity. I prefer honestly the more indigenous, where the research is led by people here on the ground, they build their own career... The condition for international collaborations here, two thirds of the funds have to be spent locally...” [MENA senior male researcher]<br>“Ideas of the projects were coming from high income countries... in low income countries we just become collectors and we would not be mentioned in articles... We put a standing rule...No research will be published in Africa without having an African contemporary as a partner.” [SSA senior male researcher] |
| language barrier/ Writing | “There is a language barrier to some extent... people have the perception that it all has to be in English and whenever it is in English then they don’t feel comfortable...” [MENA senior female researcher]<br>“We are not taught how to write.... I just have to say it... I think to have Americans or white people looking at it, they have to be polite because it can be a very sensitive thing for them to say anything. Our writing is atrocious.” [SSA senior female researcher] |
| Mentorship/<br>sponsorship/next gen | ” ...mentorship, mentorship-attitude, mentorship-growth. encouraging people to truly mentor and build the next generation... requires a little bit of selflessness... you have to be willing to allow someone else to overtake you... How do we grow that culture, In a world where everybody wants to be top dog.” [SSA senior female researcher]<br>“Structured mentoring is very important where you have senior people mentoring the students then you are also having peer to peer mentoring among the students and the post-docs. So mentoring is going top-down but also bottom up and sideways, that’s very important.” [SSA senior female researcher] |
|  | “Researcher is really an apprenticeship, its learning by apprenticeship, which is mentorship.” [SSA senior male researcher] |
| Opportunities | “as part of this building, the ecosystem or the scientific community, they even give funding for high school students to do research, opportunities for internships. They go to schools sometimes and talk about research.” [MENA senior male researcher]<br>“Unfortunately, in the region there aren’t really attractive opportunities. It’s hard to find good opportunities especially when you compare resources. It was actually a struggle to find such an opportunity.” [MENA senior male researcher] |
| Publishing | “So I have written in Lancet that we have to support our national journals because if we send our papers to national journals then they will readily accept it, welcome it and publish it... instead of running after fancy journals who are very snobbish and who are going to refuse our papers. In the world of social media, it doesn’t matter where the paper is published, once you have the DOI, you can share it on email, on twitter, on face book... It will be known to everybody that you have published this paper... it doesn't have to be New England Journal of Medicine, American Journal of Surgery, Lancet or JAMA.” [ASIA senior male researcher] |
| Research experience | “I had a really good training in different science cultures and of course part of the culture here... I think it’s really important to get this training because people get exposed to a different way of thinking, to have things really done in an established scientific program and then when they come back, they come back with that experience to use here.” [MENA senior male researcher] |
| Research for research sake | “Students reached out to me... interested in getting our internship, from hospitals and colleges based in Bahrain... the students don’t want to research... they want the benefit of research that will get attached on their C.V when they are applying to the U.S.A or UK or Europe.” [ASIA senior male researcher] |
| Training & excellence | “So how do you connect what is in your head with your heart for good. I think training... I am a living blue print of a star. It’s not that.....I am not making it up... I was trained as an undergraduate to do research and that I articulated to what I did in PhD.” [SSA senior female researcher]<br>“We are not training people in how to communicate and disseminate research especially in the written form. Oral is all very good and its excellent... but a lot of what goes to really change the research world needs to be written, needs to be published, needs to be in this peer reviewed format.” [SSA senior female researcher] |
| Trends and re-learning | “Nairobi University sits on their laurels that they are the best. Moi University came from being an underdog to trying to see what they could do, they were open to collaborations, they were open to more, they were open to opportunities ... Moi definitely has given Nairobi a run for its money.” [SSA senior female researcher] |

| RECOGNITION | Quotes |
| --- | --- |
| Self-recognition (confidence/ imposter syndrome) | “We have limitations, we have historical issues but you still have to learn and the thing is, our brains are just as set up as wired as anybody else. We have to get our minds to do the work.” [SSA senior female researcher]<br>“It is really more about the doubts that we have in ourselves you know, because you have been trained...some people will think like I am not cut out for this, I am not good enough, so it’s real that lack of self-efficacy. lack of belief. I think it is really overcoming **that internalized goliath** we should start early... We have to make people know that you have something to contribute... that can make a difference.” [SSA senior male researcher] |
| Inequity and bias (epistemic injustice) | “when I joined university, it was still very much patriarchal and like very condescending and don’t see it disappeared because there are still some people who sort of say,” you won’t be able to do that, it’s kind of impossible” So, I have taken it the other way round, every time somebody tells me you can’t do it, it’s impossible, I am like, “I am determined to show you that you are wrong,” [SSA senior female researcher]<br>“When I come home, I can do some research work or play with my children but for a woman, the same responsibilities plus running the family..., it is not taken very serious by institutions when a woman is asking for research funding and a man is different. It will be a burden on the woman” [SSA senior male researcher]<br>“Journals reject and are suspicious of African articles [paraphrased]. If I just send an article to the Lancet from Ethiopia and say your ”African name.” They will say who is “African name”? It’s very difficult, but if its someone like “John Miller” sending an article from Harvard sending, I don’t think it will have the same acceptance.” [SSA senior male researcher]<br>“Funds are there but you know, funds are...should I speak really? Funds are given to their favorites. There is a lot of favoritism... Every nation has crafty institutes, Central institutes where important people go for their treatment. The funding goes to these big institutions because they are in the funding committee of all the government departments..., it hardly ever comes to grassroots, never.” [ASIA senior male researcher] |
| Breaking glass ceilings | “But you have to break the glass ceiling. They will not offer it for you, they will not offer it for me. The global note, we have to break many glass ceilings and that is just one of them. Nobody is going to say, “Come on, welcome” Nobody is going to put a red carpet for you, come, you are welcome. You have to break each and every glass ceiling, but they will never talk about it, they will always say, “we are promoting women,” but it is not going to happen.” [ASIA senior male researcher] |
| Success definition | “if I stayed in the USA I would not have had the same success as I have it here, it’s definitely way more and the reason for this is because the resources here are better, in the USA, it is extremely competitive you know like getting a grant, it’s very competitive right now but here the competition is definitely less. So this allowed me to build much bigger things, to do lots of things yah. So, I have been very, very productive.” [MENA senior male researcher]<br>“Of course I can talk of the papers, at personal level these things really give a lot of contentment. I mean just to see a whole generation you know, coming through, getting trained, and now they are actively working in many different places and they are active researchers and they are generating papers.” [MENA senior male researcher]<br>“There needs to be high profiled status within publication groups and not like middle authorship... So for me it’s about the respect and then and the recognition of the partners in those groups.” [SSA senior male researcher] |
| Communication & Public relations | “PR for research is not good. We do not do enough story telling about our research... even lay people do not know what you are doing, so there is just not enough PR on research to let it garner the economics needed to do the work. Students do not know the value..., in the community people don’t understand it, politicians don’t even know it... You know when McDonald is coming to town; it has good PR... with beautiful colors that make people get attracted to. Beer, tobacco, all have got good PR, they call in what you want, they pick it up and people use it. Research has no PR.” [SSA senior female researcher] |
| **COLLABORATION** | ”It’s also about finding the right collaborators, building those personal relationships with the funding agents... what makes you smart is building those relationships with NIH grant program officers, the Gates foundation with the EU, the Canadians, those interpersonal relationships and networks... that’s where you hear information and things you have not seen published online.” [SSA senior male researcher]<br>“As part of this model, they contract several American universities to open campuses here and yah I got recruited fortunately in one of those universities called world Cornell medical college, which is part of the Cornell university in |
|  | the USA." [MENA senior male researcher]<br>"Nature has a deal with the Egyptian government represented in the EKB, Egyptian Knowledge Bank, where they give classes to university staff and researchers to learn how better to write papers, what are the criteria, how to edit the papers and how the reviewers see it." [MENA junior female researcher] |
| **CULTURE** | "I think I would put some of that money in changing behaviors and mental model of potential young researchers, I am not giving up on the old ones but I think I would go to the ones that are still coming up. I will put significant resources in engaging the early, just as they are choosing careers to be letting them know where research is place, whether you are doing medicine, public health, nursing even social sciences, similar thing." [SSA senior male researcher]<br>"You need to surround yourself with the people who understand. It might seem like it is work to you, but this is life and death for some of us. And so I want to be around people who value that, who appreciate that, it is not about 8-5, it is about life and death." [SSA senior female researcher] |

